# An integrative multi-context Mendelian randomization method for identifying risk genes across human tissues

**DOI:** 10.1101/2024.03.04.24303731

**Authors:** Yihao Lu, Ke Xu, Bowei Kang, Brandon L. Pierce, Fan Yang, Lin S. Chen

## Abstract

Mendelian randomization (MR) provides valuable assessments of the causal effect of exposure on outcome, yet the application of conventional MR methods for mapping risk genes encounters new challenges. One of the issues is the limited availability of expression quantitative trait loci (eQTLs) as instrumental variables (IVs), hampering the estimation of sparse causal effects. Additionally, the often context/tissue-specific eQTL effects challenge the MR assumption of consistent IV effects across eQTL and GWAS data. To address these challenges, we propose a multi-context multivariable integrative MR framework, mintMR, for mapping expression and molecular traits as joint exposures. It models the effects of molecular exposures across multiple tissues in each gene region, while simultaneously estimating across multiple gene regions. It uses eQTLs with consistent effects across more than one tissue type as IVs, improving IV consistency. A major innovation of mintMR involves employing multi-view learning methods to collectively model latent indicators of disease relevance across multiple tissues, molecular traits, and gene regions. The multi-view learning captures the major patterns of disease-relevance and uses these patterns to update the estimated tissue relevance probabilities. The proposed mintMR iterates between performing a multi-tissue MR for each gene region and joint learning the disease-relevant tissue probabilities across gene regions, improving the estimation of sparse effects across genes. We apply mintMR to evaluate the causal effects of gene expression and DNA methylation for 35 complex traits using multi-tissue QTLs as IVs. The proposed mintMR controls genome-wide inflation and offers new insights into disease mechanisms.

## Introduction

Mendelian randomization (MR) examines the causal relationships between risk exposures and complex disease outcomes, using genetic variants as instrumental variables (IVs).^1–4^ With the rapidly growing availability of summary statistics from genome-wide association studies (GWASs), two-sample MR leveraging two sets of GWAS summary statistics as input has achieved many successes in assessing the causal effects of complex traits as exposures for diseases.^5–11^ Recently, transcriptome-wide MR (TWMR) considers gene expression as risk exposure and leverages expression quantitative trait loci (eQTL) and GWAS summary statistics to map risk genes.^12–15^ Unlike transcriptome-wide association studies (TWAS),^16,17^ TWMR focuses on causal assessment. Comparing with colocalization analysis,^18–22^ MR offers the flexibility to adjust for known confounders,^23,24^ consider joint exposures,^25–28^ and allow unmeasured confounders under appropriate assumptions.^8–11^

While MR offers valuable insights, the application of conventional MR methods in TWMR analysis for mapping risk genes comes with new challenges.^12,29–32^ A notable issue is the limited number of eQTLs as IVs,^32,33^ with cis-eQTLs being generally correlated.^33^ Furthermore, disease-associated eQTLs tend to have tissue-specific effects,^34^ while the disease-relevant tissue types are often unknown.^35,36^ This can lead to inconsistent IV effects across GWAS and eQTL samples, violating core IV assumptions.^5,37,38^ These issues motivate us to consider multiple tissues simultaneously. Nevertheless, in multi-tissue MR analysis, the causal effects of genes on diseases are often tissue-specific and sparse,^39–41^ and thus the estimation of tissue-specific causal effects with a limited number of eQTLs/IVs is challenging.

Recognizing these challenges and opportunities, we propose a multi-context multivariable integrative Mendelian randomization method – mintMR, specifically designed for mapping gene expression and molecular traits as risk exposures. For each gene, we perform a multi-tissue MR analysis using eQTLs with non-zero and sign-consistent effects in more than one tissue as IVs, thereby improving the IV consistency. Our method improves the estimation of tissue-specific causal effects of all genes by simultaneously modeling the latent tissue indicators of disease relevance for multiple gene regions, jointly learning the major/low-rank patterns of latent indicators/probabilities via multi-view learning techniques, and then using the major patterns to estimate and update the probability of non-zero effects. The rationale is that risk genes for a disease often show non-zero effects in similar or related tissues,^34,36^ and by jointly learning the major patterns across genes, one can gain improved estimation of tissue-relevance probabilities and further use them to estimate the tissue-specific causal effects for each gene. The joint learning of disease-relevance of latent tissue indicators improves the estimation of sparse tissue-specific causal effects for all genes. Our algorithm iterates between estimating multi-tissue MR models for each gene and jointly learning the latent patterns and probabilities of non-zero causal effects for all genes until the maximum iteration is reached. Our MR framework considers cis gene expression and DNA methylation (DNAm) as joint exposures. Given the frequent co-occurrence of eQTLs and mQTLs,^30^ the joint consideration of DNAm with gene expression is crucial for accurately mapping causal genes. If the causal DNAm is associated with gene expression and cis-eQTLs selected as IVs are also associated with DNAm, the DNAm would be a confounder being associated with IV, and its omission could lead to biased causal inference. By jointly assessing the causal effects of gene expression and DNAm, we demonstrate that the proposed method controls genome-wide inflation, improves the power, and offers valuable insights into disease-relevant tissues and mechanisms. Our mintMR approach uniquely tackles challenges in mapping molecular traits as risk exposures via MR, jointly learns the low-rank patterns in the probabilities of disease relevance across many genes, and thereby enhances the estimation of sparse tissue-specific causal effects.

## Methods

### A starting model for a single gene region

We start with a multi-tissue MR model for studying the gene expression of a single gene from multiple tissues as the exposure and a complex disease as the outcome. We consider an eQTL *i* (*i* = 1, …, *I*_*g*_) as an IV for the expression of a gene indexed by *g*. Let *γ*_*gik*_ (*k* = 1, …, *K*) denote the true marginal effect of the SNP *i* on the gene *g* in tissue *k*. Let 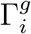 denote the true marginal association between SNP *i* and the disease outcome of interest, and the superscript *g* indicates that the SNP *i* is an IV for gene *g*. Denote 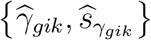 as the estimated SNP-gene association and its standard error for SNP *i* and gene *g* in tissue *k*, and 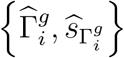 as the estimated effect of SNP *i* on the outcome and its standard error. We have the model for SNP *i*:

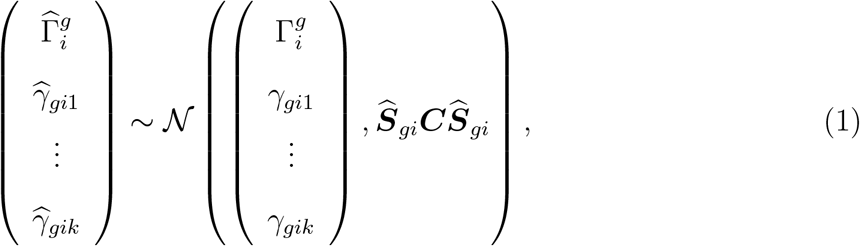

where ***C*** is the tissue-tissue correlation matrix due to sample overlap and is often estimated apriori,^42^ 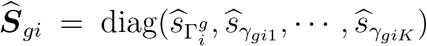 is the standard error estimate from GWAS and multi-tissue eQTL studies.

We further assume the true causal relationship between GWAS and eQTL effects, 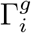 and *γ*_*gik*_’s, is linear and is given by

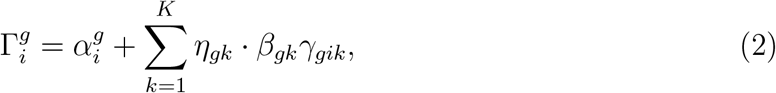

where *β*_*gk*_ is the causal effect of interest for gene *g* in tissue *k*. We introduce *η*_*gk*_ as a latent indicator for disease relevance of tissue, and *η*_*gk*_ = 1 if *β*_*gk* ≠_ 0. We assume *η*_*gk*_ ∼ Bernoulli(*π*_*gk*_). The effect of gene expression levels on the disease outcome is often sparse and varies across contexts/tissues/cell types. The effect *η*_*gk*_ · *β*_*gk*_ is the direct effect of the gene *g* in tissue *k* on the disease outcome not mediated via other exposures (including the gene expression in other tissues). When estimating the latent variables and the causal effects, the estimated probability of non-zero for the latent indicator can be viewed as a weight on the relevance of tissue types or the proportion of disease-relevant cell types in the current tissues. Without modeling latent disease-relevance tissue indicators, all tissues in the model are weighted equally. Here the true IV-to-exposure effect follows 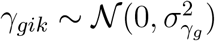 and 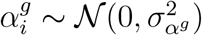 is the uncorrelated horizontal pleiotropic effect (green arrow in Figure 1a) when IV affects outcome not through exposure and IV is not associated with confounder.

**Figure 1.**
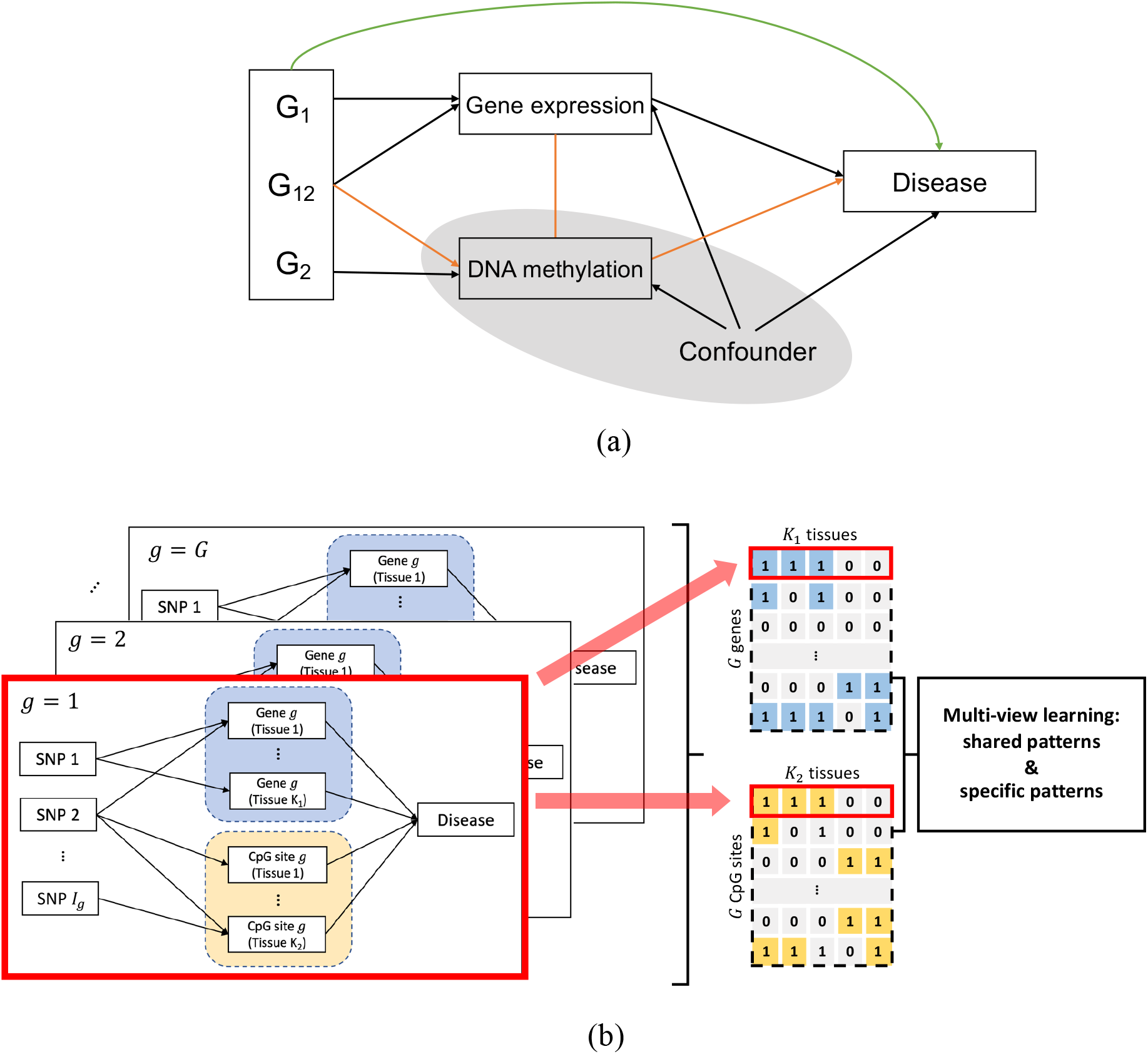
Illustrations of the multi-context multivariable integrative Mendelian randomization method. (a) The causal diagram of the multivariable MR model. When assessing the effect of gene expression on outcome, if a correlated exposure (e.g., DNA methylation; shaded) is not considered in the model, it will serve as a confounder and bias the inference (orange line). The green line represents uncorrelated horizontal pleiotropic effect. (b) An illustration of the mintMR framework for analyzing multiple gene-CpG pairs from *G* gene regions. MintMR takes as input *G × L* (*L* = 2 here) sets of IV-to-exposure effects and standard error matrices from multi-tissue eQTL and mQTL studies, respectively. It models the latent status for each causal effect. Via a logit function, mintMR links the latent status of the causal effects with a continuous modulation matrix. By performing multi-view learning on the modulation matrices, mintMR captures the low-rank data-shared and data-specific major patterns and uses them to estimate the disease-relevant probabilities. By iterating between performing MR for each gene region and estimating the disease-relevant probabilities for all genes, mintMR improves the estimation and inference on sparse causal effects for all genes.

Additionally, we consider a multivariable MR (MVMR) framework for a set of *L* (*l* = 1, …, *L*) molecular traits as exposures, each from *K*_*l*_ contexts/tissues. For example, in our motivating application, we jointly consider a gene expression and a CpG site from multiple tissues as the exposures, *L* = 2. Let SNP *i* be a cis-molecular QTL (xQTLs) for gene *g*, and 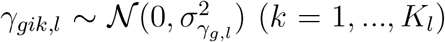 denote the marginal effect of SNP *i* on the *l*-th molecular exposure in tissue *k*. Extending model (2), we assume the following causal relationship holds between the marginal effect of the SNP *i* on the outcome, i.e., 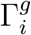, and the marginal effects of the SNP *i* on exposures, i.e.,*γ*_*gik*,*l*_’s:

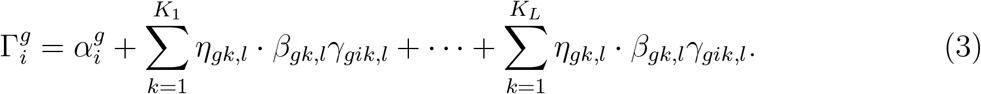

In model (3), *η*_*gk*,*l*_ · *β*_*gk*,*l*_ describes the direct effect of exposure *l* in tissue *k* on the outcome not operating through the exposure in other tissues nor through other exposures (*l*′≠ *l*). Here similar to model (2), we assume *η*_*gk*,*l*_ ∼ Bernoulli(*π*_*gk*,*l*_) and 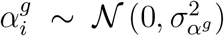 . The MVMR model allows the joint modeling of correlated cis-molecular traits in the gene regions to identify the risk factors and elucidate the mechanisms. In practice, since often there are only a limited number of xQTLs as IVs, the causal effects (and the latent indicators) in the above single-gene models (2) and (3) may not be statistically identifiable.

### The proposed mintMR model for jointly learning the disease-relevance of tissue indicators across *G* gene-CpG pairs

Common eQTLs are often weakly selected and disease-associated genetic variants typically influence downstream genes with effects being highly context-specific.^34^ When multiple genes are causally affecting diseases in a pathway or gene set, they often have effects specific to certain disease-associated tissues and cell types. Furthermore, the enrichment of disease-associated gene expression has been successfully used to identify disease-relevant tissues and cell types.^36^ These observations motivate us to jointly learn the patterns of disease-relevance indicators/probabilities across many genes, especially considering the sparse nature of disease-relevant causal effects.

We propose a joint MVMR model across *G* gene-CpG pairs to estimate the causal effects for each gene and CpG in each tissue and jointly learn the major patterns of latent disease-relevance tissue indicators, particularly in scenarios where these effects are sparse. As illustrated in Figure 1b, we consider multi-tissue expression and DNAm of the gene-CpG pairs from the *g*-th gene region (*g* = 1, · · ·, *G*) and study their effects on the outcome. While the direct effects *β*_*gk*,*l*_’s may vary in magnitude and direction, there could still be concerted patterns among the true non-zero causal effects and their effect operating contexts/tissues. The proposed mintMR model works by iteratively estimating the starting model (3) for each gene-CpG pair (one red box in Figure 1b) and collectively capturing the low-rank (major) patterns of non-zero causal effects across *G* gene regions for updating the tissue-relevance probabilities/weights until the maximum iteration is reached. The resulting estimates provide not only the causal effect for each gene and CpG site, but also the estimated probability of disease relevance for each gene-tissue or CpG-tissue pair accounting for shared patterns. A major innovation of our model is the use of multi-view learning methods to capture the low-rank patterns shared across gene regions and omics-data types. The details of the estimation are provided in Algorithm 1.

To learn the low-rank patterns of disease-relevance (non-zero causal effects) across genes, molecular exposures, and tissue types, one may employ multi-view learning strategies such as co-training,^43^ multiple kernel learning,^44^ and canonical correlation analysis (CCA).^45,46^ For each gene-CpG pair, we have model (3). We model the latent disease-relevance tissue indicators for all *G* gene-CpG-tissue trios, assuming the latent indicator *η*_*gk*,*l*_ ∼ Bernoulli(*π*_*gk*,*l*_). As illustrated in Figure 1b, we form *L* latent disease-relevance indicator matrices for *L* molecular exposures, 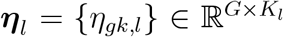 for expression and DNAm (*L* = 2) in our motivating application. We introduce a continuous modulation matrix for each exposure *l*, 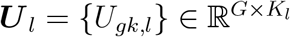, and

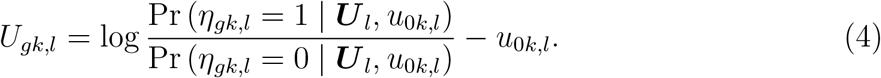

Here ***U*** _*l*_ modulates the probability of the latent binary association status, *u*_0*k*,*l*_ is the tissue-specific intercept, controlling the sparsity of non-zero effects in the *k*-th tissue of the *l*-th type of molecular exposure. For each gene (*g* = 1, · · ·, *G*), we estimate model (3) separately and then jointly model the *L* modulation matrices. We approximate the modulation matrices, ***U*** _*l*_’s, with low-rank matrices 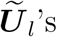 capturing the major patterns of disease-relevance across gene regions, molecular exposures, and tissue types. The mintMR model uses these approximated low-rank matrices 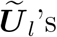 to estimate the disease-relevance probability for each gene/CpG in each tissue without over-parameterization. If there is no pattern shared across gene regions/molecular exposures/tissues, *U*_*gk*,*l*_ = 0, ∀*g, k, l*, and model (4) is reduced to logit (*π*_*gk*,*l*_) = *u*_0*k*,*l*_, i.e., only tissue-specific prior being imposed for the indicators across exposures.

More specifically, in this work, we capture the major patterns of disease relevance across all genes as the sum of major patterns shared across molecular exposures (expression and DNAm) and major tissue-sharing patterns specific to each data type. We have

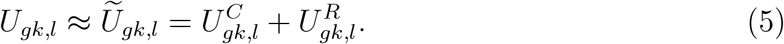

The matrices 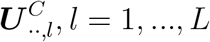 represent the common major structures shared across the *L* latent tissue-relevance indicator matrices. We estimate 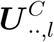 by applying generalized CCA on the matrices 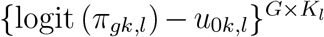. Furthermore, the 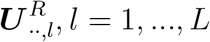 matrices capture omics data-specific tissue-sharing patterns. We perform separate principal component analysis (PCA) on each residual matrix 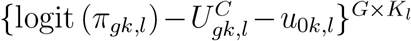 to obtain the low-rank patterns in each omics exposure data type, 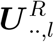 . Alternative multi-view learning methods could be used to capture different types of desirable data patterns and obtain other approximated matrices.^43–46^ The proposed mintMR algorithm iterates between estimating the causal effects in the single-gene model (3) for each of the *G* gene regions and jointly learning/estimating the latent disease-relevance indicators/probabilities via Gibbs sampling until the maximum iteration is reached (see Algorithm 1 for details).

### Accounting for LD

When studying gene expression and DNAm as joint molecular exposures, the number of e/mQTLs as IVs is generally limited. Applying a stringent LD clumping threshold would lose many IVs and hurt power. Instead of assuming independent IVs as in most existing multivariable MR methods,^47^ we allow IVs to be correlated. Assuming non-overlapping samples, we model the estimated effect sizes by accounting for the correlation among IVs *i* = 1, …, *I*_*g*_:

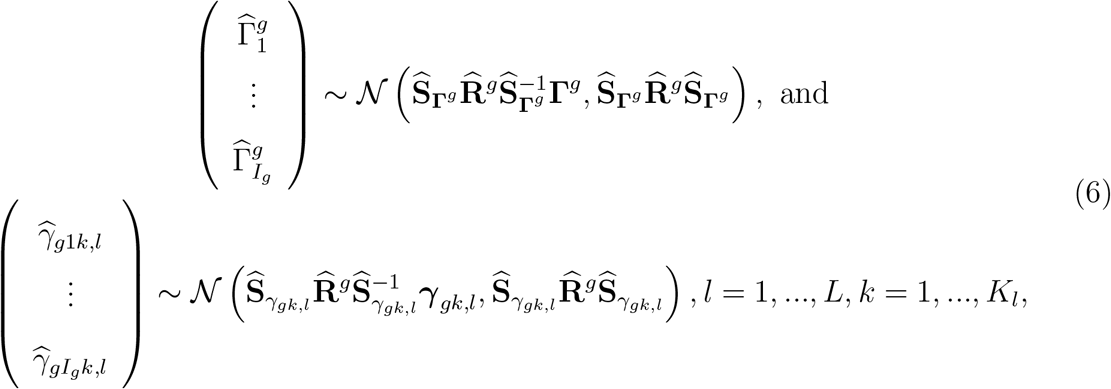

where 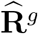 is the correlation matrix of the *I*_*g*_ number of IVs for the *g*-th set of exposures, 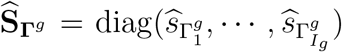 and 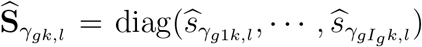. In Supplemental Information, we provided details of the Gibbs sampling algorithm of mintMR accounting for both LD and sample overlap.

## Results

### Simulations to evaluate the performance of mintMR versus competing MR methods

We conducted simulation studies to evaluate the performance of mintMR in comparison with existing univariable MR (UVMR) and MVMR methods in various scenarios.

We simulated individual-level data for the GWAS study of outcome and multi-tissue QTL studies for each exposure (details in Supplemental Information). We simulated a genotype matrix for each gene-CpG pair *g*, with all generated SNPs having uncorrelated horizontal pleiotropy (UHP) effects on the simulated outcome not via exposures. We varied the proportion of the variance in the outcome that can be explained by these UHP effects. We then generated matrices of disease-relevance tissue indicators ***η***_*l*_’s, where *η*_*gk*,*l*_ ∼ Bernoulli(*π*_*gk*,*l*_). Outcome variables in the GWAS study were simulated according to the data generation models in (S2). QTL data were simulated based on the model (S3). With generated individual-level data, we calculated the marginal QTL and GWAS summary statistics as the input for MR analyses.

Most existing MR methods were developed to analyze complex traits as exposure. In TWMR, the number of cis-eQTLs as IVs for gene expression as exposure is generally much smaller than the number of IVs in conventional MR analyses. Our simulation studies show that the limited number of IVs poses a challenge for existing MR methods. We compared mintMR with existing multivariable methods, including MVMR-IVW,^25^ MVMR-Egger,^26^ MVMR-Lasso,^27^ MVMR-Median,^27^ MVMR-Robust^27^ and MVcML.^28^ In addition, we included IVW with cross-tissue IVs and IV effects being estimated based on a meta-analysis of multiple tissue types (termed as “IVW+metaIV” below) and MR-Egger in the comparison. Among those competing methods, IVW and MVMR-IVW do not allow invalid IVs;^25,48^ MR-Egger and MVMR-Egger require Instrument Strength Independent of Direct Effect (InSIDE) assumption;^6,26^ MVMR-Median assumes the majority of IVs are valid;^27^ MVMR-Lasso and MVMR-Robust are robust to outliers (few invalid IVs);^27^ and MVcML requires plurality condition where the valid IVs form the largest group to give the causal parameter estimate.^28^ All existing UVMR and MVMR methods are developed for using complex traits as exposures. Here we adapted them to TWMR with molecular traits as exposures for comparison purposes. Moreover, we compared the proposed mintMR with its two variations: mintMR_oracle_ is a variation of mintMR where the true latent disease-relevance indicator is known, and it provides the optimal performance of mintMR, which in practice cannot be achieved without further information on disease-relevance indicators; and mintMR_single-gene_ performs the starting model (3) for each single gene region separately without the joint learning of shared patterns, and its comparison to the proposed mintMR illustrates the improvement gained by jointly learning low-rank disease-relevance patterns across multiple gene regions, tissues, and molecular exposures. We applied competing MVMR methods with multiple tissues of both simulated expression and DNA methylation as exposures and applied MR-Egger with a single tissue of gene expression as exposure to evaluate their performance. We presented the comparison of type I error rates and powers of the proposed mintMR versus competing methods at the *p*-value threshold of 0.05. To evaluate the estimation of effect sizes, we also compared the root-mean-square errors (RMSEs) of all methods (see Supplemental Information).

For each simulation, we generated *G* = 50 pairs of genes and CpGs (*L* = 2) from 5 tissues (*K*_*l*_ = 5, *l* = 1, 2), each with 500 samples. We generated 15 IVs for each gene-CpG pair and included IVs with *p*-value*<* 0.01 in at least one tissue. We simulated two types of causal effects of genes on outcomes. In the first setting (Table 1a), we simulated genes having effects on outcome in multiple tissues, with effect indicators *η*_*gk*,*l*_’s having the same probability (*π*_*gk*,*l*_ = 0.05) across all tissues. In the second setting (Table 1b), 15% of the genes have non-zero effects on outcome in one tissue (*π*_*gk*,*l*_ = 0.15). In each of the rest tissues, 3% of the genes have non-zero effects with *π*_*gk*_*′*,*l* = 0.03. We varied the proportion of the variation in the outcome explained by UHP effects of the 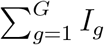 IVs of all *G* genes from 0.05 to 0.15. As shown in both Table 1a and Table 1b, when the number of IVs was limited and all IVs had UHP,^11^ the proposed mintMR model could control type I error rate. Most competing methods, including mintMR_single-gene_, suffered from inflated type I error rates. Most of the competing methods showed increases in type I error rates when UHP effects increased. MVMR-Robust had reasonable control of type I error rates but suffered from low power. When the proportion of the variation in the outcome explained by UHP effects increased, the powers of all methods decreased. The proposed mintMR method had comparable power to the oracle method, mintMR_oracle_. These simulation results, in particular the comparisons of mintMR with mintMR_oracle_ and mintMR_single-gene_, suggested that multi-view learning of shared patterns across multiple genes can effectively improve the estimation of latent disease-relevant probabilities, which leads to the improved estimation of the causal effects of interest. In Table S1, we showed that mintMR had the smallest RMSE among all the methods. In both settings, the multi-view learning of low-rank patterns of causal effects improved the power and precision when the number of IVs was limited.

**Table 1:**
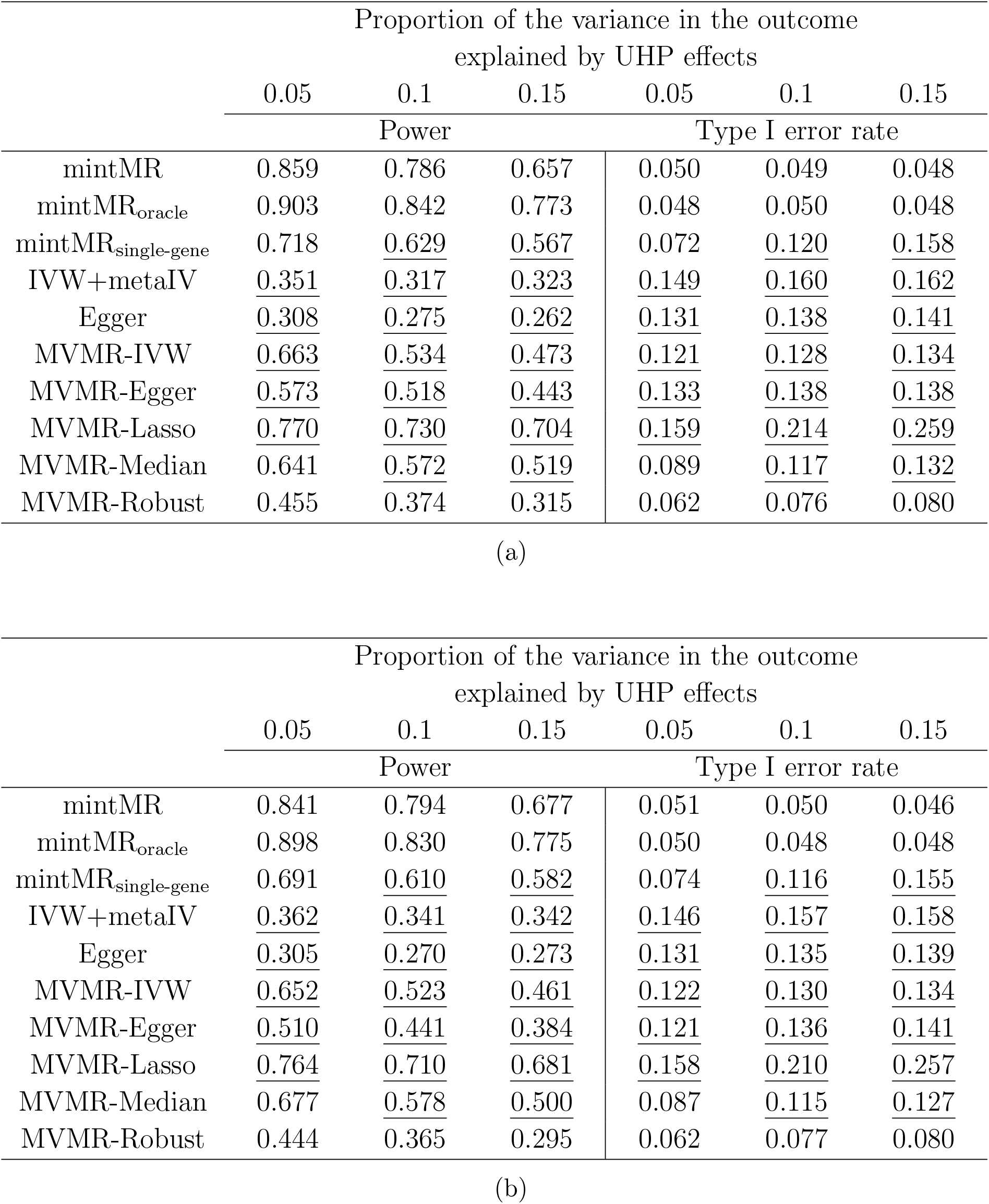
Simulation results evaluating the performance of mintMR versus competing methods when the number of IVs is limited. Two types of causal effects of genes on outcomes are simulated. (a) Genes affect outcomes in multiple tissues, with each gene having an equal probability (5%) of having non-zero effects in any tissue. (b) In one tissue, 15% of the genes have non-zero effects on outcome. In each of the rest tissues, 3% of the genes have non-zero effects. The proportion of variation in outcome explained by UHP effects varies from 0.05 to 0.15. The sample size of the outcome is 50,000 and 500 for exposure. The number of IVs is 15. Two exposures are generated and each exposure has 5 tissues. The causal effects are generated with 𝒩 (0, 0.015). Results are underlined for methods unable to control type I error rates (≥ 0.1).

In Table 2, we compared these methods in different scenarios. In Table 2a, we increased the number of IVs from 15, 25, to 100. When the number of IVs increased, all competing methods could better control the type I error rates. When the number of IVs was 100, all MVMR methods had reasonable control of the type I error rates. The power and RMSE (Table S2) of all competing MVMR methods were similar to the proposed mintMR and mintMR single-gene version. The univariable MR methods IVW and Egger still had slightly inflated type I error rates and low power due to the omission of correlated exposures. These competing methods were proposed for analyzing complex traits as exposures, and the number of IVs in conventional MR analyses is usually much larger than the number of cis-QTLs as IVs in TWMR analyses. In other words, while existing MR methods work effectively for complex trait exposures, they may not perform as well in TWMR analyses, and our proposed mintMR was tailored for analyzing molecular traits as exposures from multiple contexts/tissues. In Table 2b, we varied the probability of QTL effect sharing. When the probability decreased, eQTL/IV effects became more context/tissue-specific and the consistency of IV effects decreased. Table 2b showed that when the consistency of QTL effects across the QTL and GWAS sample decreased, power was reduced for all methods due to the inclusion of many inconsistent IVs. Conversely, the power improved when more QTLs with tissue-shared effects were selected as IVs. This simulation underscores the importance of considering multiple tissues and selecting QTLs with consistent effects across more than one tissue as IVs. In Table 2c, we varied the number of tissues for each exposure. When the number of tissues increased, mintMR showed improved power as more IVs were included.

**Table 2:** Simulation results evaluating the performance of mintMR versus competing methods. (a) tion results when varying the number of IVs. The proportion of variation in outcome explained P effect is 0.1. The causal effects are generated from 𝒩 (0, 0.01). The probability of QTL effect consistent across QTL and GWAS samples is 0.8. Five tissues are generated for each exposure. mulation results when decreasing the probability of QTL effect being consistent. The causal effects nerated from 𝒩 (0, 0.02). We simulated 15 IVs across 5 tissues for each exposure. (c) Simulation when the number of tissues increases. We simulated 15, 25, and 45 IVs when the number of for each exposure increased from 5, 10, to 20, respectively. The probability of consistency is 0.8. effects are generated from 𝒩 (0, 0.01). Results are underlined for methods unable to control type rates (≥ 0.1).

|  | Number of IVs |  |  |  |  |  |
| --- | --- | --- | --- | --- | --- | --- |
|  | 15 | 25 | 100 | 15 | 25 | 100 |
|  | Power |  |  | Type I error rate |  |  |
| mintMR | 0.734 | 0.822 | 0.932 | 0.049 | 0.055 | 0.041 |
| mintMR<sub>oracle</sub> | 0.764 | 0.861 | 0.964 | 0.049 | 0.051 | 0.056 |
| mintMR<sub>single-gene</sub> | 0.547 | 0.789 | 0.963 | 0.115 | 0.109 | 0.059 |
| IVW+metaIV | 0.351 | 0.352 | 0.530 | 0.145 | 0.144 | 0.093 |
| Egger | 0.236 | 0.254 | 0.220 | 0.134 | 0.112 | 0.074 |
| MVMR-IVW | 0.444 | 0.774 | 0.969 | 0.122 | 0.064 | 0.055 |
| MVMR-Egger | 0.383 | 0.729 | 0.898 | 0.122 | 0.063 | 0.055 |
| MVMR-Lasso | 0.631 | 0.783 | 0.969 | 0.194 | 0.068 | 0.057 |
| MVMR-Median | 0.526 | 0.745 | 0.920 | 0.114 | 0.116 | 0.100 |
| MVMR-Robust | 0.276 | 0.730 | 0.961 | 0.066 | 0.047 | 0.051 |

(a)
|  | Probability of QTL effect being consistent across QTL and GWAS sample |  |  |  |  |  |
| --- | --- | --- | --- | --- | --- | --- |
|  | 0.8 | 0.5 | 0.2 | 0.8 | 0.5 | 0.2 |
|  | Power |  |  | Type I error rate |  |  |
| mintMR | 0.867 | 0.789 | 0.660 | 0.053 | 0.066 | 0.060 |
| mintMR<sub>oracle</sub> | 0.898 | 0.852 | 0.746 | 0.049 | 0.054 | 0.050 |
| mintMR<sub>single-gene</sub> | 0.751 | 0.712 | 0.527 | 0.236 | 0.152 | 0.194 |
| IVW+metaIV | 0.299 | 0.388 | 0.399 | 0.148 | 0.156 | 0.168 |
| Egger | 0.367 | 0.289 | 0.264 | 0.109 | 0.110 | 0.127 |
| MVMR-IVW | 0.682 | 0.572 | 0.436 | 0.122 | 0.106 | 0.067 |
| MVMR-Egger | 0.562 | 0.495 | 0.407 | 0.120 | 0.098 | 0.064 |
| MVMR-Lasso | 0.793 | 0.679 | 0.443 | 0.188 | 0.129 | 0.069 |
| MVMR-Median | 0.723 | 0.671 | 0.500 | 0.172 | 0.112 | 0.069 |
| MVMR-Robust | 0.513 | 0.432 | 0.324 | 0.067 | 0.052 | 0.036 |

(b)
|  | Number of tissues for each exposure |  |  |  |  |  |
| --- | --- | --- | --- | --- | --- | --- |
|  | 5 | 10 | 20 | 5 | 10 | 20 |
|  | Power |  |  | Type I error rate |  |  |
| mintMR | 0.553 | 0.713 | 0.739 | 0.052 | 0.037 | 0.062 |
| mintMR<sub>oracle</sub> | 0.586 | 0.810 | 0.900 | 0.048 | 0.050 | 0.048 |
| mintMR<sub>single-gene</sub> | 0.497 | 0.538 | 0.583 | 0.174 | 0.235 | 0.126 |
| IVW+metaIV | 0.321 | 0.286 | 0.281 | 0.146 | 0.157 | 0.125 |
| Egger | 0.240 | 0.180 | 0.192 | 0.138 | 0.124 | 0.105 |
| MVMR-IVW | 0.316 | 0.302 | 0.487 | 0.126 | 0.122 | 0.081 |
| MVMR-Egger | 0.266 | 0.326 | 0.404 | 0.126 | 0.153 | 0.084 |
| MVMR-Lasso | 0.612 | 0.364 | 0.493 | 0.303 | 0.145 | 0.081 |
| MVMR-Median | 0.418 | 0.225 | 0.480 | 0.135 | 0.076 | 0.103 |
| MVMR-Robust | 0.175 | 0.190 | 0.407 | 0.072 | 0.062 | 0.060 |

We presented additional simulation results in Supplemental Information. In Table S3, we showed that mintMR had the smallest RMSEs when varying the consistency of QTL effect and the number of tissues. In Table S4, we increased the sample size from 500 to 10,000 for each tissue type in the presence of UHP. The larger tissue sample size improved the estimation of the IV-to-exposure effects, while also making the impacts of invalid IVs stronger. The performance of competing methods was similar for different sample sizes. In Table S5, we varied the causal effect size, the proposed mintMR method controlled the type I error rate and showed improved power compared with other methods. MintMR had the smallest RMSEs on data with varied sample sizes and effect sizes (Table S6). In addition, we simulated correlated IVs with genetic correlation up to 0.5. When the IVs were correlated and the numbers of IVs were limited, the proposed mintMR could still control the type I error rate and showed reasonable power (Table S7).

### Data analysis: Identifying trait/disease risk-associated genes via mintMR

We applied the proposed mintMR method to map risk genes for 35 complex traits and diseases, including 14 immunological traits, 6 metabolic traits, 2 neurological diseases, 2 cardiovascular traits, 7 psychiatric diseases and traits, and 4 other traits. We used GWAS statistics as the IV-to-outcome statistics. Details of the GWAS statistics can be found in Table S11. We used multi-tissue eQTL and mQTL summary statistics as the IV-to-exposure statistics. For eQTLs, we obtained the summary statistics for blood tissue from the eQTLGen consortium^49^ (*N* = 31, 684), for muscle tissue (*N* = 706), lung tissue (*N* = 515) and brain cerebellum tissue (*N* = 209) from version 8 of the Genotype-Tissue Expressions (GTEx) project,^33^ and for brain dorsolateral prefrontal cortex tissue from the Religious Orders Study and Memory and Aging Project (ROSMAP; *N* = 560).^50^ For mQTLs, we obtained the summary statistics for lung tissue from GTEx^18^ (*N* = 190), skeletal muscle tissue from FUSION^51^ (*N* = 265), and blood tissue (*N* = 1, 366) from Brisbane Systems Genetics Study (BSGS)^52,53^ plus Lothian Birth Cohorts (LBC).^54^ For each gene, we selected the proximal CpG site (within 100 KB of TSS) that explained the most variation in expression. For each gene-CpG pair, we selected the cis-eSNPs or mSNPs with non-zero and sign-consistent eQTL or mQTL effects in at least two tissues (*P* ≤ 0.005). We performed LD clumping at the *r*^2^ threshold of 0.01. We restricted our analysis to genes with at least 10 IVs overall and at least one IV for each tissue.

We applied mintMR to each of the 35 complex traits and diseases, with an average of 3,440 genes examined for each trait/disease. At the false discovery rate (FDR) of 0.05, we identified the genes and CpG sites showing significant effects in at least two tissues for each examined trait/disease. See Table S8 for a list of examined traits/diseases, the number of genes studied, and the number of detected genes and CpG sites. In Table S9, we evaluated the genome-wide inflation factor^55^ with and without accounting for DNAm, based on the *p*-value distributions of gene expression in each tissue. By accounting for the most correlated cis-CpG site, genome-wide inflation is substantially reduced for all examined traits and diseases. An important message from our analysis result is that in mapping the expression of risk genes, cis-DNAm can be a major confounder if not accounted for. Existing studies showed that cis DNAm frequently correlates with cis expression and cis-eQTLs often co-occur with cis-mQTLs^30^. If a cis-e/mQTL or a variant in LD with it is selected as an IV and cis-DNAm is not accounted for, the causal inference can be compromised due to the IVs being correlated with the confounder. In Table S10, we showed that mintMR had lower inflation factors than MVMR-Lasso, MVMR-Median, and MVMR-IVW. The inflation factors of mintMR and MVMR-Robust are comparable. Due to the prevalent pleiotropy in TWMR analysis, MVMR-Egger and MVMR-Robust are expected to have lower power than the other examined methods.^27,28^ Simulation showed that MVMR-Egger and MVMR-Robust have much lower power than mintMR when UHP is prevalent. We also note that there is remaining mild inflation in the *p*-values. It suggests that there are additional factors and potential IV-associated confounders that have not been fully accounted for in the analyses. This could be at least partially due to, for example, secondary cis-CpG sites, and other correlated and co-expressed cis genes in the region. The proposed mintMR model is a multivariable MR framework and it can be applied to jointly consider one or more cis gene expression and multiple CpG sites.

In Figure 2a, we showed the quantile-quantile (QQ) plot of negative log base 10 of *p*-values for gene expression effects on hypertension in the blood tissue. The genome-wide inflation factor decreased from 1.88 to 1.25 after accounting for DNAm. In the 5q31-32 region, we identified four genes (*HSPA4, HARS2, KIAA0141*, and *ARHGEF37*) showing significant effects on hypertension (FDR*<* 0.05) without accounting for DNAm. After adjusting for the most correlated cis-CpG site, only the expression of *HSPA4* still showed a significant effect (Figure 2b). *HSPA4* is a member of the heat shock protein 70 family, which is known to be involved in the pathogenesis of hypertension.^56,57^ We further conducted a colocalization analysis, and only the gene *HSPA4* showed a high probability of colocalization with hypertension (PP4= 0.95) (Figure 2c). Additionally, we examined all the significant genes identified for hypertension at the FDR level of 0.05 in at least two tissues. Out of the 57 identified genes, 49 were analyzed in a TWAS^58^. Among these, 15 genes (30.6%) were also significant in the TWAS analysis (*P <* 0.005), a proportion much higher than that observed among all genes examined (14.2%). Moreover, 6 out of these 49 genes (12.2%) were supported by colocalization analyses (PP4*>* 0.7), a much higher proportion than all genes examined (2.3%).

**Figure 2.**
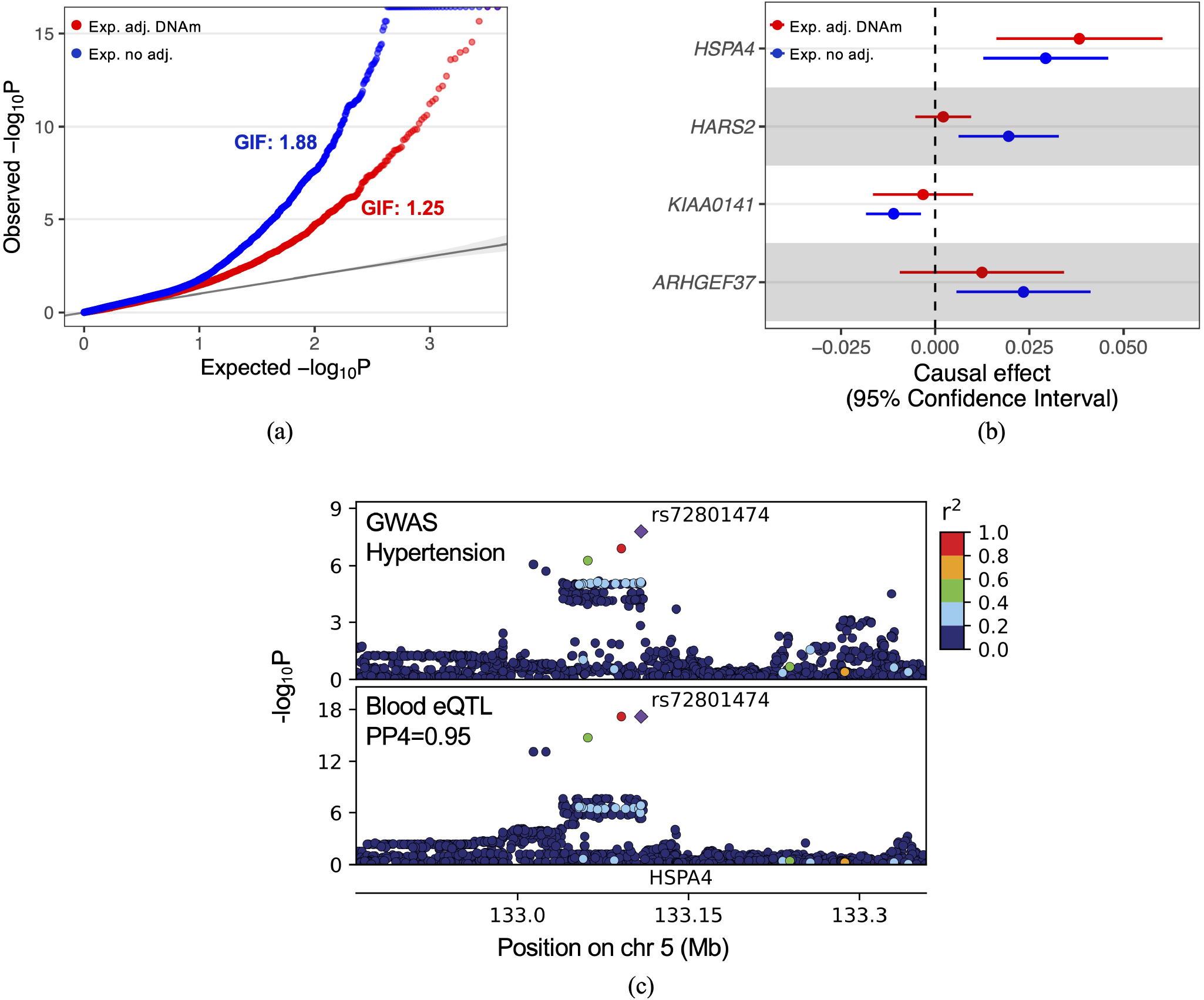
(a) A QQ plot of negative log base 10 of *p*-values for gene expression effects on hypertension. Red points represent the *p*-values of gene expression adjusting for DNAm. Blue points are the *p*-values of gene expression without adjusting for DNAm. Genome-wide inflation factors (GIF) for both analyses are shown. (b) The causal effects of four genes on hypertension in the blood tissue in the 5q31-32 region, with and without adjusting for DNAm. Without adjusting for DNAm (blue points and error bars), the four gene expression levels show significant effects on hypertension (FDR*<*0.05). After adjusting for DNAm (red points and error bars), only the expression of *HSPA4* is significant. (c) Genotype-phenotype association *p*-values in the *HSPA4* locus for hypertension GWAS (top panel) and eQTL in the blood (bottom panel). The colocalization probability (PP4) of eQTL with GWAS signal is shown. The diamond-shaped point represents the top significant eQTL variant (rs72801474). Linkage disequilibrium between SNPs is assessed by squared Pearson coefficient of correlation (*r*^2^).

We further conducted pathway analyses on the significant genes and proximal genes correlated with significant CpGs identified for each of the 35 traits and diseases, utilizing the Reactome^59^ and Gene Ontology^60^ database. We detected the significantly enriched biological pathways for each trait/disease, as shown in Figure 3. Our results revealed many enriched pathways being shared among related traits, suggesting shared mechanisms. Lipid-related pathways, including lipid localization and transport, are implied for Alzheimer’s disease, monocyte count lymphocyte count, and platelet count. As the basic component of cell membranes, lipids play an important role in brain function. Impaired homeostasis of lipids is known to be related to neurologic disorders.^61–63^ Monocytes, lymphocytes, and platelets are key components of the immune system,^64–66^ and the fact that these traits share common enriched pathways with Alzheimer’s disease suggests that inflammation and immune response play a significant role in Alzheimer’s disease.^67,68^

**Figure 3.**
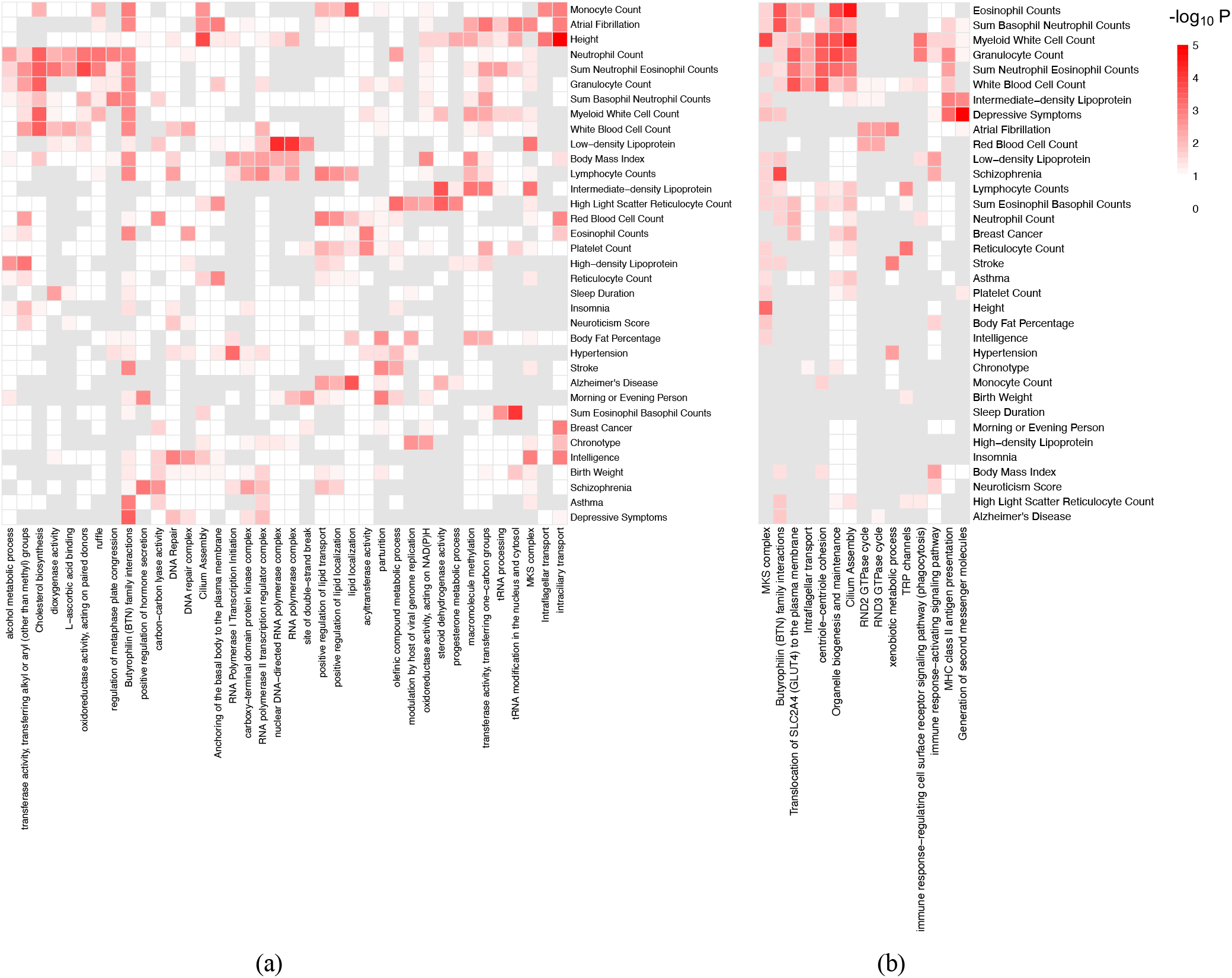
The heatmaps of enriched pathways for (a) identified genes affecting complex traits/diseases and (b) proximal genes correlated with the identified significant CpG sites. The *p*-values of pathway enrichment are calculated based on one-sided Fisher’s exact tests without multiple testing adjustments. Pathways with *p*-values*<* 0.005 for at least two traits are presented.

## Discussion

In this work, we propose an integrative multi-context Mendelian randomization method, mintMR, for addressing unique challenges in TWMR analysis. MintMR performs a multi-tissue MR analysis using QTLs as IVs for each gene region. It improves the estimation of tissue-specific causal effects of all genes by simultaneously modeling the latent disease-relevance context/tissue indicators for multiple gene regions, jointly learning the low-rank patterns of latent indicators/probabilities via multi-view learning techniques, and then using the major patterns to update the probability of non-zero effects. The joint learning of disease-relevance of latent tissue indicators improves the estimation of sparse tissue-specific causal effects for all genes. By selecting cross-tissue QTLs as IVs and considering both gene expression and DNAm as joint exposures, mintMR improves IV consistency and reduces confounding due to correlated cis molecular traits when mapping causal genes. Simulations show that mintMR can control the type I error rates and has good powers in various settings, even when there are a limited number of QTLs as IVs and the causal effects are sparse.

We applied mintMR to map risk genes for 35 complex traits and diseases, leveraging QTL summary statistics from multiple tissues of different studies and GWAS summary statistics. Our results showed a reasonable control of genome-wide inflation for the examined traits and diseases, demonstrating the feasibility of leveraging multitissue QTLs and jointly learning disease-relevance probabilities across multiple gene regions in improving causal identification. Our results also suggested DNAm being a major confounder in mapping risk genes. By accounting for cis DNAm, genome-wide inflation for TWMR analyses was substantially reduced. Our analysis and results demonstrated that mintMR could offer valuable insights into disease-relevant tissues and the underlying mechanisms.

There are several limitations of our work. First, mintMR does not allow IV to be associated with unmeasured confounders. As a multivariable MR framework, mintMR allows the adjustment and joint modeling of correlated molecular traits (confounders) as joint exposures. Simulation studies show that mintMR is robust to mild violations of the InSIDE assumption. In the TWMR analysis of 35 traits and diseases, we noted some remaining mild genome-wide inflation after modeling the most correlated cis-CpG sites. In future analyses, additional correlated cis molecular traits, such as secondary cis CpG sites or nearly co-expressed genes, could also be modeled to further reduce genome-wide inflation. Second, we assume linear effects of exposures on outcome. The current mintMR model is not flexible for modeling complex interactions among exposures and interactions with known covariates, such as sex-biased effects.

In future work, mintMR can be extended to allow for correlated horizontal pleiotropy by identifying IVs with such effects. Another area of future development is to improve the modeling of major patterns of disease relevance indicators by adopting other advanced multi-view learning techniques. In this work, we used CCA and PCA to capture omics-shared and tissue-shared patterns in mapping risk genes. Other deep learning and supervised multi-view learning methods could be implemented to promote other desirable patterns among examined genes.^45,69,70^ Moreover, the mintMR model could be further expanded to model interaction effects among joint exposures and covariates.

These developments will be explored in future works.

## Data Availability

All data produced in the present work are contained in the manuscript.

## Data and code availability

All the summary statistics used in this paper are publicly available. They can be accessed via the corresponding references. The code for mintMR is available at https://github.com/ylustat/mintMR.

## Acknowledgements

We thank the GTEx Consortium. The research of L.S.C. and Y.L. was supported by NIH 2R01GM108711, R35ES028379 and 1R01CA229618. Y.L. was also supported by Susan G. Komen^®^ TREND21675016.

## Author contributions

L.S.C. conceived the project. L.S.C., F.Y., and Y.L. developed the methods and wrote the manuscript. K.X. assisted Y.L. with the development of the estimation algorithm. Y.L. conducted the simulations and analyzed the data. All authors provided valuable suggestions for the development of the methods and the data analyses. All authors reviewed and approved the final manuscript.

## Declaration of interests

The authors declare no competing interests.

### Algorithm 1

The Gibbs sampling algorithm for mintMR model

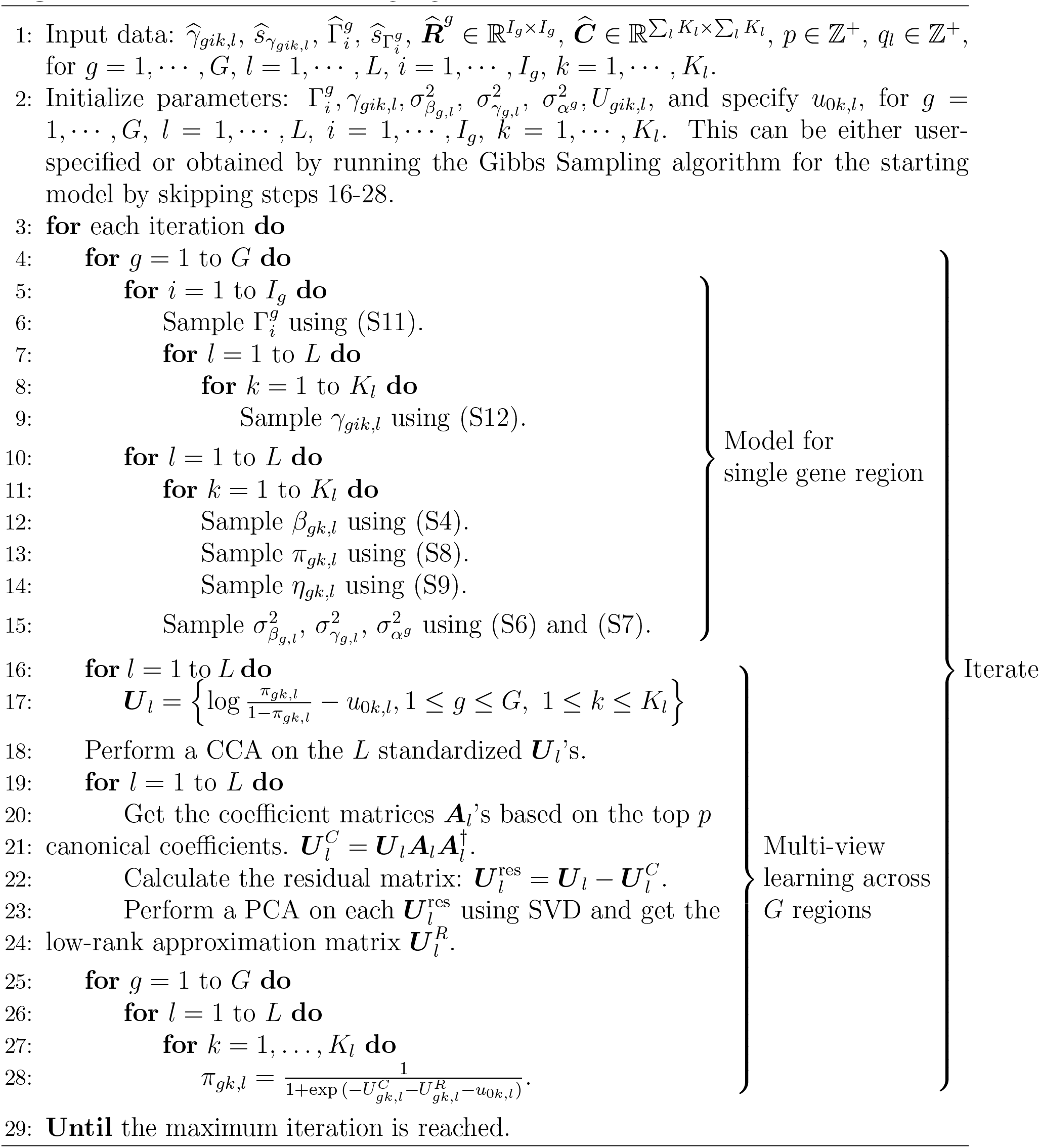

