## Supplemental Information for "An integrative multi-context Mendelian randomization method for identifying risk genes across human tissues"

### 1 Simulation Studies

#### 1.1 Data generation

In the simulation studies discussed in the main text, we simulated individual-level data for $N^y$ individuals in the GWAS study of outcome and $N^x_{gk,l}$ individuals for tissue $k$ of exposure $l$ in the multi-tissue QTL studies. In most simulations, we set $N^y = 50,000$ and $N^x_{gk,l} = 500$. We simulated an $N^y \times I_g$ genotype matrix $\boldsymbol{Q}_g$ for each gene-CpG pair $g$ with $I_g = 15$. The minor allele frequency (MAF) of each SNP follows Unif$(0.05, 0.5)$. The correlation between SNPs $i$ and $j$ is $r_{ij} = r^{|i-j|}$, where $r = 0$ in most simulations. To be consistent with the prevalent pleiotropy in TWMR analysis,[1] all generated SNPs have a direct effect on the complex trait not via gene expression or DNA methylation. We also generated a $G \times K_l$ matrix of binary indicators $\boldsymbol{\eta}_l$ for each exposure $l$, where $\eta_{gk,l} \sim$ Bernoulli$(\pi_{gk,l})$. We simulated the outcome in the GWAS study according to the following data generation models:

$$X_{gk,l} = \boldsymbol{Q}_g \boldsymbol{\mu}^x_{gk,l} + \mu^{z_x}_{gk,l} Z_g + \varepsilon_{gk,l}, \tag{S1}$$

$$Y = \sum_{g=1}^{G} \sum_{l=1}^{L} \sum_{k=1}^{K_l} \eta_{gk,l} \cdot \beta_{gk,l} X_{gk,l} + \sum_{g=1}^{G} \boldsymbol{Q}_g \boldsymbol{\mu}^y_g + \sum_{g=1}^{G} \mu^{z_y}_g Z_g + \varepsilon. \tag{S2}$$

In model (S1), the vector $X_{gk,l}$, of length $N^y$, corresponds to the values of exposure $l$ in tissue $k$ for the gene-CpG pair indexed by $g$. $\boldsymbol{\mu}^x_{gk,l} = [\mu^x_{g1k,l}, \cdots, \mu^x_{gI_g k,l}]^\top$ is the QTL effect of eSNPs. For the $i$-th SNP, we sampled $\boldsymbol{\mu}^x_{gi\cdot,\cdot} = [\mu^x_{gi1,1}, \cdots, \mu^x_{giK_L,L}]^\top$ from $\mathcal{N}(0, \boldsymbol{\Sigma}_{\mu_x})$, where the diagonal elements of $\boldsymbol{\Sigma}_{\mu_x}$ are set to 0.3 and the off-diagonal elements are fixed at 0.03, indicating a correlation coefficient of 0.1 across exposures and tissues. $Z_g$ is a vector of a latent confounder sampled from $\mathcal{N}(0, 1)$; $\mu^{z_x}_{gk,l} \sim \mathcal{N}(0, 0.1)$ is the effect of the confounder on exposures; $\varepsilon_{gk,l}$ is the error term sampled from $\mathcal{N}(0, \sigma^2_{e_x})$, with errors from different exposures and tissues being correlated with a coefficient of 0.1. In model (S2), $Y$ is a vector of a continuous trait; $\beta_{gk,l} \sim \mathcal{N}(0, \sigma^2_\beta)$ is the effect of exposure $l$ in tissue $k$ of the $g$-th gene-CpG pair on the outcome; $\eta_{gk,l}$ is an binary effect indicator following Bernoulli$(\pi_{gk,l})$; $\boldsymbol{\mu}^y_g \sim \mathcal{N}(0, \boldsymbol{\Sigma}_{\mu_y})$ is the vector of direct effects of SNPs on $Y$, with $\mathbf{\Sigma}_{\mu_y}$ being a diagonal matrix; $\mu_g^{z_y} \sim \mathcal{N}(0, 0.1)$ is the effect of the confounder on the outcome; and $\varepsilon \sim \mathcal{N}(0, \sigma^2_{e_y})$.

We generated the QTL data based on the following model:

$$\widetilde{X}_{gk,l} = \widetilde{\boldsymbol{Q}}_g \left( \boldsymbol{\mu}^x_{gk,l} \circ \boldsymbol{\delta}_{gk,l} \right) + \mu^{z_x}_{gk,l} \widetilde{Z}_g + \widetilde{\varepsilon}_{gk,l}, \tag{S3}$$

where $\widetilde{X}_{gk,l}$ is a vector of length $N^x_{gk,l}$, representing the value of exposure $l$ in tissue $k$ from the QTL study; $\widetilde{\boldsymbol{Q}}_g$ is a $N^x_{gk,l} \times I_g$ genotype matrix of the $I_g$ eSNPs in the QTL study; $\boldsymbol{\delta}_{gk,l}$ is a vector of $I_g$ Bernoulli variables for eSNPs, where $\delta_{gk,l} = 1$ indicates that the eSNP shares consistent QTL effects between the $k$-th tissue of the QTL study and the GWAS study. In most simulations, $\Pr(\delta_{gk,l} = 1) = 0.8$. The operator $\circ$ is the Hadamard product operator. With the simulated individual-level data, we calculated the marginal QTL and GWAS summary statistics as the input for the MR analyses.

#### 1.2 Additional results

As discussed in the main text, we simulated two types of causal effects of genes on outcomes. In the first setting, the effect indicators $\eta_{gk,l}$'s were simulated based on the same probability ($\pi_{gk,l} = 0.05$) across all tissues. In the second setting, we simulated a higher proportion of the causal genes having effects on outcomes in one tissue ($\pi_{gk,l} = 0.15$). In each of the rest tissues, 3% of the genes have non-zero effects with $\pi_{gk',l} = 0.03$. In Table S1, we showed that all methods had increased RMSEs when the proportion of the variance in the outcome explained by UHP effects increased. MintMR had the lowest RMSEs in both settings. Compared to mintMR$_{\text{single-gene}}$, mintMR's RMSEs were substantially lower, highlighting the improvement in effect estimation with a limited number of IVs via the multi-view learning process. In the following simulations, we simulated the effect indicators $\eta_{gk,l}$'s with the same probability ($\pi_{gk,l} = 0.05$) across all tissues.

As shown in Table S2, all methods demonstrated improved estimation performance when the number of IVs increased. In particular, when the number of IVs was large

(# IVs=100), all methods showed comparable RMSEs. MintMR also outperformed other methods in terms of RMSEs for varied numbers of analyzed tissues and varied probabilities of QTL effect sharing (Table S3). Specifically, when varying the number of analyzed tissues, we set the probability of QTL effect sharing to 0.8. When varying the probability of QTL effect sharing, each exposure had 5 tissues. Moreover, we varied the sample size from 500 to 10,000 for each tissue type (Table S4). With a larger sample size, MVcML had a substantially improved power while controlling the type I error rate. With varying causal effect sizes in Table S5, mintMR controlled the type I error rate and showed improved power compared to other methods. In the first set of simulations shown in Table S6, we varied the sample sizes of exposures from 500 to 10,000, with causal effects generated from $\mathcal{N}(0, 0.02)$. In the second set of simulations shown in Table S6, we varied the causal effect sizes, with the sample size of exposure fixed at 500. MintMR achieved the lowest RMSEs compared to competing methods. Furthermore, in the simulations with correlated IVs (Table S7), mintMR was able to control the type I error rate effectively with limited numbers of IVs and increased genetic correlation.

### 2 The Gibbs sampling algorithms for mintMR

In this section, we provide the details of the Gibbs sampler used in the mintMR estimation algorithm.

#### 2.1 The algorithm for independent SNPs

For the $g$-th set of exposures, we propose the following Bayesian hierarchical model for independent SNPs and non-overlapping samples.

$$\widehat{\gamma}_{gik,l} \mid \gamma_{gik,l}, \widehat{s}^2_{\Gamma_i^g} \sim \mathcal{N}(\gamma_{gik,l}, \widehat{s}^2_{\gamma_{gik,l}}),\ \widehat{\Gamma}_i^g \mid \Gamma_i^g, \widehat{s}^2_{\Gamma_i^g} \sim \mathcal{N}(\Gamma_i^g, \widehat{s}^2_{\Gamma_i^g}),$$

$$\gamma_{gik,l} \left| \sigma^2_{\gamma_{g,l}} \sim \mathcal{N}\left(0, \sigma^2_{\gamma_{g,l}}\right), \quad \Gamma_i^g \right| \beta_{g\cdot,\cdot}, \gamma_{gi\cdot,\cdot}, \eta_{g\cdot,\cdot}, \sigma^2_{\alpha^g} \sim \mathcal{N}\left(\sum_{l=1}^{L}\sum_{k=1}^{K_l} \eta_{gk,l}\beta_{gk,l}\gamma_{gik,l}, \sigma^2_{\alpha^g}\right),$$

$$\beta_{gk,l} \mid \sigma^2_{\beta_{g,l}} \sim \mathcal{N}\left(0, \sigma^2_{\beta_{g,l}}\right), \quad \alpha_i^g \mid \sigma^2_{\alpha^g} \sim \mathcal{N}\left(0, \sigma^2_{\alpha^g}\right)$$

$$\sigma^2_{\beta_{g,l}} \sim \mathcal{IG}\left(a_\beta, b_\beta\right), \quad \sigma^2_{\gamma_{g,l}} \sim \mathcal{IG}\left(a_\gamma, b_\gamma\right), \quad \sigma^2_{\alpha^g} \sim \mathcal{IG}\left(a_\alpha, b_\alpha\right),$$

$$\eta_{gk,l} \mid \pi_{gk,l} \sim \pi_{gk,l}^{\eta_{gk,l}}(1-\pi_{gk,l})^{1-\eta_{gk,l}}, \quad \pi_{gk,l} \sim \mathrm{Beta}(a_\pi, b_\pi),$$

where $i = 1, 2\cdots, I_g$, $k = 1, 2\cdots, K_l$, and $l = 1, \cdots, L$.

Denote $\widehat{\mathbf{\Gamma}}^g = [\widehat{\Gamma}_1^g, \cdots, \widehat{\Gamma}_{I_g}^g]^\top$, $\widehat{\boldsymbol{\gamma}}_{gk,l} = [\widehat{\gamma}_{g1k,l}, \cdots, \widehat{\gamma}_{gI_gk,l}]^\top$, $\mathbf{\Gamma}^g = [\Gamma_1^g, \cdots, \Gamma_{I_g}^g]^\top$, and $\boldsymbol{\gamma}_{gk,l} = [\gamma_{g1k,l}, \cdots, \gamma_{gI_gk,l}]^\top$, the posterior likelihood is in the form of

$$\begin{aligned}
L(\Theta_g) \propto& \prod_{g=1}^{G} p\left(\mathbf{\Gamma}^g, \boldsymbol{\gamma}_{g1,1}, \cdots, \boldsymbol{\gamma}_{gK_L,L}, \boldsymbol{\beta}_g, \sigma^2_{\mathbf{\Gamma}^g}, \sigma^2_{\alpha^g}, \boldsymbol{\eta}_g, \boldsymbol{\pi}_g \mid \widehat{\mathbf{\Gamma}}^g, \widehat{\boldsymbol{\gamma}}_{g\cdot,\cdot}\right) \\
=& \prod_{g=1}^{G}\left\{\prod_{i=1}^{I_g}\left[p(\widehat{\Gamma}_i^g | \alpha_i^g + \sum_{l=1}^{L}\sum_{k=1}^{K_l} \beta_{gk,l}\eta_{gk,l}\gamma_{gik,l}, \widehat{s}_{\Gamma_i^g}) \prod_{l=1}^{L}\prod_{k=1}^{K_l} p(\widehat{\gamma}_{gik,l} | \gamma_{gik,l}, \widehat{s}_{\gamma_{gik,l}}) p(\gamma_{gik,l} \mid \sigma^2_{\gamma_{g,l}})\right]\right\} \\
& \left\{\prod_{g=1}^{G} p(\sigma^2_{\alpha_g}) \prod_{i=1}^{I_g} p(\alpha_i^g \mid \sigma^2_{\alpha_g})\right\} \left\{\prod_{g=1}^{G}\prod_{l=1}^{L} p(\sigma^2_{\gamma_{g,l}})\right\} \\
& \left\{\prod_{g=1}^{G}\prod_{l=1}^{L} p(\sigma^2_{\beta_{g,l}}) \prod_{k=1}^{K_l} p(\beta_{gk,l} \mid \sigma^2_{\beta_{g,l}})\right\} \left\{\prod_{g=1}^{G}\prod_{l=1}^{L}\prod_{k=1}^{K_l} p(\eta_{gk,l} \mid \pi_{gk,l}) p(\pi_{gk,l})\right\}.
\end{aligned}$$

Here $p(\widehat{\Gamma}_i^g | \alpha_i^g + \sum_{l=1}^{L}\sum_{k=1}^{K_l} \beta_{gk,l}\eta_{gk,l}\gamma_{gik,l}, \widehat{s}_{\Gamma_i^g}) = p(\widehat{\Gamma}_i^g | \Gamma_i^g, \widehat{s}_{\Gamma_i^g})$.

The conditional posterior distribution of each $\Gamma_i^g$ given the other parameters in the model is

$$\Gamma_i^g \mid \widehat{\Gamma}_i^g,\ \widehat{s}_{\Gamma_i^g},\ \gamma_{gi\cdot,\cdot},\ \beta_{g\cdot,\cdot},\ \eta_{g\cdot,\cdot},\ \sigma^2_{\alpha^g} \sim \mathcal{N}\left(\widetilde{\mu}_{gi0}, \widetilde{\sigma}^2_{gi0}\right),$$

where

$$-\frac{1}{2\widetilde{\sigma}^2_{gi0}} = -\frac{1}{2}\left(\frac{1}{\widehat{s}^2_{\Gamma_i^g}} + \frac{1}{\sigma^2_{\alpha^g}}\right),$$

$$\frac{\widetilde{\mu}_{gi0}}{\widetilde{\sigma}^2_{gi0}} = \frac{\widehat{\Gamma}_i^g}{\widehat{s}^2_{\Gamma_i^g}} + \frac{\sum_{l=1}^{L}\sum_{k=1}^{K_l}\eta_{gk,l}\beta_{gk,l}\gamma_{gik,l}}{\sigma^2_{\alpha^g}}.$$

The conditional distribution for each element $\gamma_{gik,l}$ comes from a normal distribution with

$$\gamma_{gik,l} \mid \widehat{\gamma}_{gik,l},\ \widehat{s}_{\gamma_{gik,l}},\ \Gamma_i^g,\ \gamma_{gi\cdot,\cdot},\ \beta_{g\cdot,\cdot}, \eta_{g\cdot,\cdot},\ \sigma^2_{\gamma_{g,l}},\ \sigma^2_{\alpha^g} \sim \mathcal{N}\left(\widetilde{\mu}_{gik,l}, \widetilde{\sigma}^2_{gik,l}\right),$$

where

$$-\frac{1}{2\widetilde{\sigma}^2_{gik,l}} = -\frac{1}{2}\left(\frac{1}{\widehat{s}^2_{\gamma_{gik,l}}} + \frac{\eta_{gk,l}\beta^2_{gk,l}}{\sigma^2_{\alpha^g}} + \frac{1}{\sigma^2_{\gamma_{g,l}}}\right),$$

$$\frac{\widetilde{\mu}_{gik,l}}{\widetilde{\sigma}^2_{gik,l}} = \frac{\widehat{\gamma}_{gik,l}}{\widehat{s}^2_{\gamma_{gik,l}}} + \frac{\eta_{gk,l}\beta_{gk,l}\left(\Gamma_i^g - \sum_{(k',l')\neq(k,l)}\beta_{gk',l'}\eta_{gk',l'}\gamma_{gik',l'}\right)}{\sigma^2_{\alpha^g}}.$$

The conditional posterior distributions of $\beta_{gk,l}$ are from normal distributions,

$$\beta_{gk,l} \mid \Gamma_i^g,\ \gamma_{gi\cdot,\cdot},\ \eta_{g\cdot,\cdot},\ \sigma^2_{\alpha^g},\ \sigma^2_{\beta_{g,l}} \sim (1-\eta_{gk,l})\mathcal{N}\left(0, \sigma^2_{\beta_{g,l}}\right) + \eta_{gk,l}\mathcal{N}\left(\mu_{\beta_{gk,l}}, \sigma^2_{\beta_{gk,l}}\right), \tag{S4}$$

where

$$-\frac{1}{2\sigma^2_{\beta_{gk,l}}} = -\frac{1}{2}\left(\frac{\eta_{gk,l}\sum_{i=1}^{I_g}\gamma^2_{gik,l}}{\sigma^2_{\alpha^g}} + \frac{1}{\sigma^2_{\beta_{g,l}}}\right),$$

$$\frac{\mu_{\beta_{gk,l}}}{\sigma^2_{\beta_{gk,l}}} = \frac{\sum_{i=1}^{I_g}\left(\Gamma_i^g - \sum_{(k',l')\neq(k,l)}\beta_{gk',l'}\eta_{gk',l'}\gamma_{gik',l'}\right)\gamma_{gik,l}}{\sigma^2_{\alpha^g}}.$$

Conditioning on the data and the other parameters in the model, the conditional posterior distribution of $\sigma^2_{\beta_{g,l}}$ is inverse-gamma,

$$\sigma^2_{\beta_{g,l}} \mid \beta_{g\cdot,l}, a_\beta, b_\beta \sim \mathcal{IG}\left(a_\beta + \frac{K_l}{2}, b_\beta + \frac{1}{2}\sum_{k=1}^{K_l}\beta^2_{gk,l}\right). \tag{S5}$$

The conditional posterior distribution of $\sigma^2_{\gamma_{g,l}}$ is also inverse-gamma,

$$\sigma^2_{\gamma_{g,l}} \mid \gamma_{g\cdot\cdot,l}, a_\gamma, b_\gamma \sim \mathcal{IG}\left(a_\gamma + \frac{I_g K_l}{2}, b_\gamma + \frac{1}{2}\sum_{k=1}^{K_l} \boldsymbol{\gamma}_{gk,l}^\top \boldsymbol{\gamma}_{gk,l}\right). \tag{S6}$$

The conditional posterior distribution of $\sigma^2_{\alpha^g}$ is also inverse-gamma,

$$\begin{aligned}&\sigma^2_{\alpha^g} \mid \Gamma^g_{\cdot}, \gamma_{g\cdot\cdot,\cdot},\ \eta_{g\cdot,\cdot},\ \beta_{g\cdot,\cdot}, a_\alpha, b_\alpha \\ \sim&\mathcal{IG}\left(a_\alpha + \frac{I_g}{2}, b_\alpha + \frac{1}{2}\left(\boldsymbol{\Gamma}^g - \sum_{l=1}^{L}\sum_{k=1}^{K_l} \eta_{gk,l}\beta_{gk,l}\boldsymbol{\gamma}_{gk,l}\right)^\top \left(\boldsymbol{\Gamma}^g - \sum_{l=1}^{L}\sum_{k=1}^{K_l} \eta_{gk,l}\beta_{gk,l}\boldsymbol{\gamma}_{gk,l}\right)\right).\end{aligned} \tag{S7}$$

The conditional posterior of $\pi_{gk,l}$ is a Beta distribution:

$$\pi_{gk,l} \mid \eta_{gk,l}, a_\pi, b_\pi \sim \text{Beta}\left(a_\pi + \eta_{gk,l}, b_\pi + 1 - \eta_{gk,l}\right). \tag{S8}$$

The conditional probability of $\boldsymbol{\eta}_{g,l}$ given $\boldsymbol{\Gamma}^g$ can be written using Bayes' theorem:

$$\Pr\left(\eta_{gk,l} = 1 \mid \boldsymbol{\Gamma}^g\right) = \frac{\Pr\left(\eta_{gk,l} = 1\right) p\left(\boldsymbol{\Gamma}^g \mid \eta_{gk,l} = 1\right)}{\Pr\left(\eta_{gk,l} = 0\right) p\left(\boldsymbol{\Gamma}^g \mid \eta_{gk,l} = 0\right) + \Pr\left(\eta_{gk,l} = 1\right) p\left(\boldsymbol{\Gamma}^g \mid \eta_{gk,l} = 1\right)}, \tag{S9}$$

where

$$\begin{aligned}&p\left(\boldsymbol{\Gamma}^g \mid \eta_{gk,l} = 1\right) \\ =&\frac{1}{\sigma_{\alpha^g}\sqrt{2\pi}} \exp\left\{-\frac{1}{2\sigma^2_{\alpha^g}}\left(\boldsymbol{\Gamma}^g - \beta_{gk,l}\boldsymbol{\gamma}_{gk,l} - \sum_{(k',l')\neq(k,l)} \eta_{gk',l'}\beta_{gk',l'}\boldsymbol{\gamma}_{gk',l'}\right)^\top \left(\boldsymbol{\Gamma}^g - \beta_{gk,l}\boldsymbol{\gamma}_{gk,l} - \sum_{(k',l')\neq(k,l)} \eta_{gk',l'}\beta_{gk',l'}\boldsymbol{\gamma}_{gk',l'}\right)\right\}, \\ &p\left(\boldsymbol{\Gamma}^g \mid \eta_{gk,l} = 0\right) \\ =&\frac{1}{\sigma_{\alpha^g}\sqrt{2\pi}} \exp\left\{-\frac{1}{2\sigma^2_{\alpha^g}}\left(\boldsymbol{\Gamma}^g - \sum_{(k',l')\neq(k,l)} \eta_{gk',l'}\beta_{gk',l'}\boldsymbol{\gamma}_{gk',l'}\right)^\top \left(\boldsymbol{\Gamma}^g - \sum_{(k',l')\neq(k,l)} \eta_{gk',l'}\beta_{gk',l'}\boldsymbol{\gamma}_{gk',l'}\right)\right\}.\end{aligned}$$

For the $l$-th exposure, we apply the CCA (when $L = 2$) or GCCA (when $L > 2$) to the standardized modulation matrices $\boldsymbol{U}_l = \{\text{logit}(\pi_{gk,l}) - u_{0k,l}\}^{G\times K_l}$, $l = 1, \cdots, L$. CCA/GCCA aims to maximize the pair-wise correlation between linear combinations of $\boldsymbol{U}_l$'s. Suppose we have rank-$p$ approximation for $\boldsymbol{U}_l$, the corresponding canonical weight matrices $\boldsymbol{A}_l = [\boldsymbol{a}_l^1, \ldots, \boldsymbol{a}_l^p]$ can be estimated. Then, for each data type $l$, the estimated low-rank matrix $\boldsymbol{U}_l^C = \boldsymbol{U}_l \boldsymbol{A}_l \boldsymbol{A}_l^{\dagger}$ with $\operatorname{rank}\left(\boldsymbol{U}_l^C\right) = p$, where $^{\dagger}$ refers to the Moore-Penrose pseudo-inverse.

To further capture the low-rank patterns in each molecular exposure data type, we perform a PCA on the residual matrix after substracting $\boldsymbol{U}_l^C$ from $\boldsymbol{U}_l$. Specifically, we first standardize the $\boldsymbol{U}_l^{\text{res}}$'s with mean zero and unit variance. We calculate a truncated Singular Value Decomposition (SVD) and keep the top $q_l$ largest singular values to approximate $\boldsymbol{U}_l^{\text{res}}$. The rank-$q_l$ approximation of $\boldsymbol{U}_l^{\text{res}}$ is denoted as $\boldsymbol{U}_l^R$. $\boldsymbol{U}_l^R$'s capture the association patterns shared among and specific to different cellular contexts (tissues). Accounting for omic-shared $\left(\boldsymbol{U}_l^C\right)$ and tissue-shared $\left(\boldsymbol{U}_l^R\right)$ patterns, we further update $\pi_{gk,l}$'s with $\pi_{gk,l} = 1/\left(1 + \exp\left(-U_{gk,l}^C - U_{gk,l}^R - u_{0k,l}\right)\right)$ (Algorithm 1).

#### 2.2 The algorithm for correlated SNPs

For correlated SNPs, we consider the following model for the $g$-th set of exposures:

$$\begin{aligned} \widehat{\boldsymbol{\Gamma}}^g &\sim \mathcal{N}\left(\widehat{\mathbf{S}}_{\boldsymbol{\Gamma}^g} \widehat{\mathbf{R}}^g \widehat{\mathbf{S}}_{\boldsymbol{\Gamma}^g}^{-1} \boldsymbol{\Gamma}^g, \widehat{\mathbf{S}}_{\boldsymbol{\Gamma}^g} \widehat{\mathbf{R}}^g \widehat{\mathbf{S}}_{\boldsymbol{\Gamma}^g}\right), \\ \widehat{\boldsymbol{\gamma}}_{gk,l} &\sim \mathcal{N}\left(\widehat{\mathbf{S}}_{\gamma_{gk,l}} \widehat{\mathbf{R}}^g \widehat{\mathbf{S}}_{\gamma_{gk,l}}^{-1} \boldsymbol{\gamma}_{gk,l}, \widehat{\mathbf{S}}_{\gamma_{gk,l}} \widehat{\mathbf{R}}^g \widehat{\mathbf{S}}_{\gamma_{gk,l}}\right), \end{aligned} \tag{S10}$$

where $\widehat{\mathbf{R}}^g$ is the correlation matrix of the $I_g$ number of IVs for the $g$-th set of exposures, $\widehat{\boldsymbol{\Gamma}}^g = [\widehat{\Gamma}_1^g, \cdots, \widehat{\Gamma}_{I_g}^g]^\top$, $\widehat{\boldsymbol{\gamma}}_{gk,l} = [\widehat{\gamma}_{g1k,l}, \cdots, \widehat{\gamma}_{gI_gk,l}]^\top$, $\widehat{\mathbf{S}}_{\boldsymbol{\Gamma}^g} = \operatorname{diag}(\widehat{s}_{\Gamma_1^g}, \cdots, \widehat{s}_{\Gamma_{I_g}^g})$, and $\widehat{\mathbf{S}}_{\gamma_{gk,l}} = \operatorname{diag}(\widehat{s}_{\gamma_{g1k,l}}, \cdots, \widehat{s}_{\gamma_{gI_gk,l}})$. The conditional posterior distribution of each $\Gamma_i^g$ given the other parameters in the model is

$$\Gamma_i^g \mid \widehat{\Gamma}_i^g,\ \widehat{s}_{\Gamma_i^g},\ \gamma_{gi\cdot,\cdot},\ \beta_{g\cdot,\cdot},\ \eta_{g\cdot,\cdot},\ \sigma_{\alpha^g}^2,\ \widehat{R}_{\cdot\cdot}^g \sim \mathcal{N}\left(\widetilde{\mu}_{gi0}, \widetilde{\sigma}_{gi0}^2\right),$$

where

$$\begin{aligned} -\frac{1}{2\widetilde{\sigma}_{gi0}^2} &= -\frac{1}{2}\left(\frac{1}{\widehat{s}_{\Gamma_i^g}^2} + \frac{1}{\sigma_{\alpha^g}^2}\right), \\ \frac{\widetilde{\mu}_{gi0}}{\widetilde{\sigma}_{gi0}^2} &= \frac{\widehat{\Gamma}_i^g}{\widehat{s}_{\Gamma_i^g}^2} - \sum_{j \neq i}\left(\frac{\widehat{R}_{ij}^g \Gamma_j^g}{\widehat{s}_{\Gamma_j^g}}\right)\frac{1}{\widehat{s}_{\Gamma_i^g}} + \frac{\sum_{l=1}^{L}\sum_{k=1}^{K_l} \eta_{gk,l}\beta_{gk,l}\gamma_{gik,l}}{\sigma_{\alpha^g}^2}. \end{aligned}$$

Here $\widehat{\mathbf{R}}^g$ is the estimated correlation matrix among all selected IVs of exposure set $g$. Conditioning on other parameters, the distribution for each element $\gamma_{gik,l}$ comes from a normal distributions with

$$\gamma_{gik,l} \mid \widehat{\gamma}_{gik,l},\ \widehat{s}_{\gamma_{gik,l}},\ \Gamma_i^g,\ \gamma_{g\cdot k,l},\ \beta_{g\cdot,\cdot}, \eta_{g\cdot,\cdot},\ \sigma^2_{\gamma_{g,l}},\ \sigma^2_{\alpha^g},\ \widehat{R}^g_{\cdot\cdot} \sim \mathcal{N}\left(\widetilde{\mu}_{gik,l}, \widetilde{\sigma}^2_{gik,l}\right),$$

where

$$-\frac{1}{2\widetilde{\sigma}^2_{gik,l}} = -\frac{1}{2}\left(\frac{1}{\widehat{s}^2_{\gamma_{gik,l}}} + \frac{\eta_{gk,l}\beta^2_{gk,l}}{\sigma^2_{\alpha^g}} + \frac{1}{\sigma^2_{\gamma_g}}\right),$$

$$\frac{\widetilde{\mu}_{gik,l}}{\widetilde{\sigma}^2_{gik,l}} = \frac{\widehat{\gamma}_{gik,l}}{\widehat{s}^2_{\gamma_{gik,l}}} - \sum_{j\neq i}\left(\frac{\widehat{R}^g_{ij}\gamma_{gjk,l}}{\widehat{s}_{\gamma_{gjk,l}}}\right)\frac{1}{\widehat{s}_{\gamma_{gik,l}}} + \frac{\eta_{gk,l}\beta_{gk,l}\left(\Gamma_i^g - \sum_{(k',l')\neq(k,l)}\beta_{gk',l'}\eta_{gk',l'}\gamma_{gik',l'}\right)}{\sigma^2_{\alpha^g}}.$$

The updates for the remaining parameters are the same as in (S4)-(S8).

#### 2.3 The algorithm accounting for sample overlap

We further consider sample overlap among tissues, molecular traits and complex traits. We could rewrite the distribution for the summary statistics in (S10) as the following matrix normal distribution for $Z$-score

$$\left[\widehat{\mathbf{S}}^{-1}_{\mathbf{\Gamma}^g}\widehat{\mathbf{\Gamma}}^g,\ \underbrace{\widehat{\mathbf{S}}^{-1}_{\boldsymbol{\gamma}_{g1,1}}\widehat{\boldsymbol{\gamma}}_{g1,1},\ \cdots,\ \widehat{\mathbf{S}}^{-1}_{\boldsymbol{\gamma}_{gK_1,1}}\widehat{\boldsymbol{\gamma}}_{gK_1,1}}_{\text{Exposure 1, context 1-}K_1},\cdots,\ \underbrace{\widehat{\mathbf{S}}^{-1}_{\boldsymbol{\gamma}_{g1,L}}\widehat{\boldsymbol{\gamma}}_{g1,L},\ \cdots,\ \widehat{\mathbf{S}}^{-1}_{\boldsymbol{\gamma}_{gK_L,L}}\widehat{\boldsymbol{\gamma}}_{gK_L,L}}_{\text{Exposure L, context 1-}K_L}\right]$$
$$\sim\mathcal{MN}\left(\left[\widehat{\mathbf{R}}^g\widehat{\mathbf{S}}^{-1}_{\mathbf{\Gamma}^g}\mathbf{\Gamma}^g,\ \widehat{\mathbf{R}}^g\widehat{\mathbf{S}}^{-1}_{\boldsymbol{\gamma}_{g1,1}}\boldsymbol{\gamma}_{g1,1},\ \cdots,\ \widehat{\mathbf{R}}^g\widehat{\mathbf{S}}^{-1}_{\boldsymbol{\gamma}_{gK_L,L}}\boldsymbol{\gamma}_{gK_L,L}\right], \widehat{\mathbf{R}}^g,\ \widehat{\boldsymbol{C}}\right),$$

where $\widehat{\boldsymbol{C}} \in \mathbb{R}^{(1+\sum_{l=1}^L K_l)\times(1+\sum_{l=1}^L K_l)}$ is the correlation matrix that accounts for sample overlap among outcome and the $\sum_{l=1}^L K_l$ contexts of the $L$ exposures. The matrix $\widehat{\boldsymbol{C}}$ can be estimated separately using summary statistics among independent variants with no associations to either exposure or outcome diseases/traits. Equivalently, it can be written as a multivariate normal distribution as:

$$\begin{pmatrix} \widehat{\boldsymbol{\Gamma}}^g \\ \widehat{\boldsymbol{\gamma}}_{g1,1} \\ \vdots \\ \widehat{\boldsymbol{\gamma}}_{gK_L,L} \end{pmatrix} \sim \mathcal{N}\left( \begin{pmatrix} \widehat{\mathbf{S}}_{\boldsymbol{\Gamma}^g} \widehat{\mathbf{R}}^g \widehat{\mathbf{S}}_{\boldsymbol{\Gamma}^g}^{-1} \boldsymbol{\Gamma}^g \\ \widehat{\mathbf{S}}_{\boldsymbol{\gamma}_{g1,1}} \widehat{\mathbf{R}}^g \widehat{\mathbf{S}}_{\boldsymbol{\gamma}_{g1,1}}^{-1} \boldsymbol{\gamma}_{g1,1} \\ \vdots \\ \widehat{\mathbf{S}}_{\boldsymbol{\gamma}_{gK_L,L}} \widehat{\mathbf{R}}^g \widehat{\mathbf{S}}_{\boldsymbol{\gamma}_{gK_L,L}}^{-1} \boldsymbol{\gamma}_{gK_L,L} \end{pmatrix}, \begin{pmatrix} \widehat{\mathbf{S}}_{\boldsymbol{\Gamma}^g} & \mathbf{0} & \cdots & \mathbf{0} \\ \mathbf{0} & \widehat{\mathbf{S}}_{\boldsymbol{\gamma}_{g1,1}} & \cdots & \mathbf{0} \\ \vdots & \vdots & \ddots & \vdots \\ \mathbf{0} & \mathbf{0} & \cdots & \widehat{\mathbf{S}}_{\boldsymbol{\gamma}_{gK_L,L}} \end{pmatrix} \left(\widehat{\boldsymbol{C}} \otimes \widehat{\mathbf{R}}^g\right) \begin{pmatrix} \widehat{\mathbf{S}}_{\boldsymbol{\Gamma}^g} & \mathbf{0} & \cdots & \mathbf{0} \\ \mathbf{0} & \widehat{\mathbf{S}}_{\boldsymbol{\gamma}_{g1,1}} & \cdots & \mathbf{0} \\ \vdots & \vdots & \ddots & \vdots \\ \mathbf{0} & \mathbf{0} & \cdots & \widehat{\mathbf{S}}_{\boldsymbol{\gamma}_{gK_L,L}} \end{pmatrix} \right).$$

We denote $\boldsymbol{\Lambda} = \{\lambda_{ij}\} = \widehat{\boldsymbol{C}}^{-1}$ $(i, j = 0, 1, \cdots, \sum_{l=1}^{L} K_l)$. For simplicity, in following equations we use $\lambda_{0k,l} = \lambda_{0\left(k+\sum_{t<l} K_t\right)}$. It represents the value in $\boldsymbol{\Lambda}$ for the sample overlap between the outcome and the $k$-th context of the $l$-th exposure. We use $\lambda_{kk',ll'} = \lambda_{\left(k+\sum_{t<l} K_t\right)\left(k'+\sum_{t<l'} K_t\right)}$. It represents the value in $\boldsymbol{\Lambda}$ for the sample overlap between the $k$-th context of the $l$-th exposure and the $k'$-th context of the $l'$-th exposure. Through some derivations, the conditional posterior distribution of each $\Gamma_i^g$ given the other parameters in the model is

$$\Gamma_i^g \mid \widehat{\Gamma}_{\cdot}^g,\ \widehat{\gamma}_{g\cdot\cdot,\cdot},\ \gamma_{g\cdot\cdot,\cdot},\ \widehat{s}_{\Gamma_{\cdot}^g},\ \widehat{s}_{\gamma_{g\cdot\cdot,\cdot}},\ \beta_{g\cdot,\cdot},\ \eta_{g\cdot,\cdot},\ \sigma_{\alpha^g}^2,\ \widehat{R}_{\cdot\cdot}^g,\ \lambda_{\cdot\cdot,\cdot\cdot} \sim \mathcal{N}\left(\widetilde{\mu}_{gi0}, \widetilde{\sigma}_{gi0}^2\right), \tag{S11}$$

where

$$\begin{aligned} -\frac{1}{2\widetilde{\sigma}_{gi0}^2} =& -\frac{1}{2}\left(\frac{\lambda_{00}}{\widehat{s}_{\Gamma_i^g}^2} + \frac{1}{\sigma_{\alpha^g}^2}\right) \\ \frac{\widetilde{\mu}_{gi0}}{\widetilde{\sigma}_{gi0}^2} =& \lambda_{00}\left\{\frac{\widehat{\Gamma}_i^g}{\widehat{s}_{\Gamma_i^g}^2} - \sum_{j\neq i}\left(\frac{\widehat{R}_{ij}^g \Gamma_j^g}{\widehat{s}_{\Gamma_j^g}}\right)\frac{1}{\widehat{s}_{\Gamma_i^g}}\right\} + \sum_{l=1}^{L}\sum_{k=1}^{K_l} \frac{\widehat{\gamma}_{gik,l}}{\widehat{s}_{\gamma_{gik,l}}} \cdot \frac{\lambda_{0k,l}}{\widehat{s}_{\Gamma_i^g}} \\ & - \sum_{l=1}^{L}\sum_{k=1}^{K_l}\sum_{j=1}^{I_g} \frac{\widehat{R}_{ij}^g \gamma_{gjk,l}}{\widehat{s}_{\gamma_{gjk,l}}} \cdot \frac{\lambda_{0k,l}}{\widehat{s}_{\Gamma_i^g}} + \frac{\sum_{l=1}^{L}\sum_{k=1}^{K_l} \eta_{gk,l}\beta_{gk,l}\gamma_{gik,l}}{\sigma_{\alpha^g}^2}. \end{aligned}$$

The conditional distribution for each element $\gamma_{gik,l}$ comes from a normal distribution with

$$\gamma_{gik,l} \mid \widehat{\Gamma}_{\cdot}^g,\ \Gamma_{\cdot}^g,\ \widehat{\gamma}_{g\cdot\cdot,\cdot},\ \widehat{s}_{\Gamma_{\cdot}^g},\ \widehat{s}_{\gamma_{g\cdot\cdot,\cdot}},\ \beta_{g\cdot,\cdot}, \eta_{g\cdot,\cdot},\ \sigma_{\alpha^g}^2,\ \widehat{R}_{\cdot\cdot}^g,\ \lambda_{\cdot\cdot,\cdot\cdot} \sim \mathcal{N}\left(\widetilde{\mu}_{gik,l}, \widetilde{\sigma}_{gik,l}^2\right), \tag{S12}$$

where

$$
\begin{aligned}
-\frac{1}{2\widetilde{\sigma}^2_{gik,l}} =& -\frac{1}{2}\left(\frac{\lambda_{kk,ll}}{\widehat{s}^2_{\gamma_{gik,l}}} + \frac{\eta_{gk,l}\beta^2_{gk,l}}{\sigma^2_{\alpha^g}} + \frac{1}{\sigma^2_{\gamma_{g,l}}}\right),\\
\frac{\widetilde{\mu}_{gik,l}}{\widetilde{\sigma}^2_{gik,l}} =& \frac{\widehat{\Gamma}^g_i}{\widehat{s}_{\Gamma^g_i}} \cdot \frac{\lambda_{0k,l}}{\widehat{s}_{\gamma_{gik,l}}} + \sum_{l'=1}^{L}\sum_{k'=1}^{K_l} \frac{\widehat{\gamma}_{gik',l'}}{\widehat{s}_{\gamma_{gik',l'}}} \cdot \frac{\lambda_{kk',ll'}}{\widehat{s}_{\gamma_{gik,l}}}\\
& - \sum_{j=1}^{I_g} \frac{\widehat{R}^g_{ij}\Gamma^g_j}{\widehat{s}_{\Gamma^g_j}} \cdot \frac{\lambda_{0k,l}}{\widehat{s}_{\gamma_{gik,l}}} - \sum_{j\neq i}\left(\frac{\widehat{R}^g_{ij}\gamma_{gjk,l}}{\widehat{s}_{\gamma_{gjk,l}}}\right)\frac{\lambda_{kk,ll}}{\widehat{s}_{\gamma_{gik,l}}} - \sum_{(k',l')\neq(k,l)}\sum_{i'=1}^{I_g} \frac{\widehat{R}^g_{ii'}\gamma_{gi'k',l'}}{\widehat{s}_{\gamma_{gi'k',l'}}} \cdot \frac{\lambda_{kk',ll'}}{\widehat{s}_{\gamma_{gik,l}}}\\
& + \frac{\eta_{gk,l}\beta_{gk,l}\left(\Gamma^g_i - \sum_{(k',l')\neq(k,l)} \beta_{gk',l'}\eta_{gk',l'}\gamma_{gik',l'}\right)}{\sigma^2_{\alpha^g}}.
\end{aligned}
$$

The updates for the remaining parameters are the same as in (S4)-(S8).

| | Proportion of the variance in the outcome explained by UHP effects | | |
|---|---|---|---|
| | 0.05 | 0.1 | 0.15 |
| mintMR | 0.027 | 0.033 | 0.038 |
| mintMR (oracle) | 0.011 | 0.012 | 0.013 |
| mintMR (single gene) | 0.081 | 0.092 | 0.107 |
| IVW+metaIV | 0.047 | 0.053 | 0.057 |
| Egger | 0.163 | 0.187 | 0.208 |
| MVMR-IVW | 0.044 | 0.055 | 0.065 |
| MVMR-Egger | 0.061 | 0.078 | 0.091 |
| MVMR-Lasso | 0.050 | 0.065 | 0.077 |
| MVMR-Median | 0.051 | 0.065 | 0.076 |
| MVMR-Robust | 0.044 | 0.055 | 0.065 |
| MVcML | 0.034 | 0.038 | 0.040 |

(a)

| | Proportion of the variance in the outcome explained by UHP effects | | |
|---|---|---|---|
| | 0.05 | 0.1 | 0.15 |
| mintMR | 0.027 | 0.033 | 0.038 |
| mintMR (oracle) | 0.011 | 0.012 | 0.014 |
| mintMR (single gene) | 0.077 | 0.089 | 0.103 |
| IVW+metaIV | 0.047 | 0.052 | 0.057 |
| Egger | 0.160 | 0.184 | 0.204 |
| MVMR-IVW | 0.044 | 0.056 | 0.065 |
| MVMR-Egger | 0.066 | 0.084 | 0.099 |
| MVMR-Lasso | 0.050 | 0.065 | 0.077 |
| MVMR-Median | 0.051 | 0.065 | 0.076 |
| MVMR-Robust | 0.044 | 0.056 | 0.065 |
| MVcML | 0.034 | 0.038 | 0.040 |

(b)

Table S1: RMSE comparison of mintMR versus its variants and competing methods when IVs are limited ($I_g = 15$). Two types of causal effects of genes on outcomes are simulated. (a) Genes affect outcomes in multiple tissues, with each gene having an equal probability (5%) of having non-zero effects in any tissue. (b) In one tissue, 15% of the genes have non-zero effects on outcome. In each of the rest tissues, 3% of the genes have non-zero effects. The proportion of the variance in the outcome explained by UHP effect varies from 0.05 to 0.15. The sample size for the outcome is 50,000 and 500 for each exposure. Two exposures are generated and each exposure has 5 tissues. The causal effects are generated from $\mathcal{N}(0, 0.015)$.

| | Number of IVs | | | | | |
|---|---|---|---|---|---|---|
| | 15 | 25 | 100 | 15 | 25 | 100 |
| | Proportion of the variance in the outcome explained by UHP effects | | | | | |
| | 0.05 | | | 0.15 | | |
| mintMR | 0.032 | 0.025 | 0.017 | 0.037 | 0.029 | 0.018 |
| $\text{mintMR}_{\text{oracle}}$ | 0.015 | 0.013 | 0.012 | 0.017 | 0.015 | 0.013 |
| $\text{mintMR}_{\text{single-gene}}$ | 0.072 | 0.051 | 0.034 | 0.090 | 0.062 | 0.036 |
| IVW+metaIV | 0.048 | 0.046 | 0.028 | 0.053 | 0.051 | 0.030 |
| Egger | 0.178 | 0.142 | 0.138 | 0.202 | 0.158 | 0.148 |
| MVMR-IVW | 0.055 | 0.029 | 0.017 | 0.065 | 0.033 | 0.018 |
| MVMR-Egger | 0.088 | 0.033 | 0.019 | 0.104 | 0.038 | 0.020 |
| MVMR-Lasso | 0.063 | 0.029 | 0.017 | 0.076 | 0.034 | 0.018 |
| MVMR-Median | 0.063 | 0.034 | 0.020 | 0.074 | 0.040 | 0.021 |
| MVMR-Robust | 0.055 | 0.029 | 0.017 | 0.065 | 0.033 | 0.018 |
| MVcML | 0.036 | 0.031 | 0.022 | 0.037 | 0.034 | 0.025 |

Table S2: RMSE comparison of mintMR versus its variants and competing methods when varying the number of IVs. Two exposures are generated and each exposure has 5 tissues. The total number of IVs varies from 15 to 100. The sample size for the outcome is 50,000 and 500 for each exposure. The causal effects are simulated from $\mathcal{N}(0, 0.01)$.

| | Number of tissues | | | Probability of QTL effect being consistent | | |
|---|---|---|---|---|---|---|
| | 5 | 10 | 15 | 0.8 | 0.5 | 0.2 |
| mintMR | 0.041 | 0.034 | 0.029 | 0.032 | 0.036 | 0.041 |
| $\text{mintMR}_{\text{oracle}}$ | 0.019 | 0.013 | 0.013 | 0.014 | 0.016 | 0.019 |
| $\text{mintMR}_{\text{single-gene}}$ | 0.106 | 0.073 | 0.051 | 0.093 | 0.080 | 0.076 |
| IVW+metaIV | 0.058 | 0.048 | 0.036 | 0.055 | 0.064 | 0.065 |
| Egger | 0.223 | 0.199 | 0.207 | 0.142 | 0.179 | 0.671 |
| MVMR-IVW | 0.073 | 0.072 | 0.040 | 0.058 | 0.060 | 0.052 |
| MVMR-Egger | 0.117 | 0.091 | 0.043 | 0.072 | 0.072 | 0.060 |
| MVMR-Lasso | 0.088 | 0.079 | 0.040 | 0.065 | 0.063 | 0.053 |
| MVMR-Median | 0.083 | 0.080 | 0.047 | 0.065 | 0.067 | 0.061 |
| MVMR-Robust | 0.073 | 0.072 | 0.040 | 0.058 | 0.060 | 0.052 |
| MVcML | 0.037 | 0.020 | 0.013 | 0.052 | 0.052 | 0.042 |

Table S3: RMSE comparison of mintMR versus its variants and competing methods when varying the number of tissues and the probability of having consistent effects in QTL and GWAS data for each IV. Specifically, when varying the number of tissues for each exposure from 5, 10, to 15, we generated 15, 25, and 45 IVs respectively, with the probability of consistent effects fixed at 0.8. When varying the probability of consistent effects from 0.8 to 0.2, we fixed the number of tissues for each exposure as 5. In both sets of simulations, the causal effects are generated from $\mathcal{N}(0, 0.02)$ and the proportion of the variance in the outcome explained by UHP effects is 0.1.

| | Number of samples of exposures | | | | | |
|---|---|---|---|---|---|---|
| | 500 | 1000 | 10000 | 500 | 1000 | 10000 |
| | Power | | | Type I error rate | | |
| mintMR | 0.833 | 0.868 | 0.872 | 0.052 | 0.052 | 0.049 |
| $\text{mintMR}_{\text{oracle}}$ | 0.878 | 0.880 | 0.922 | 0.045 | 0.049 | 0.052 |
| $\text{mintMR}_{\text{single-gene}}$ | 0.707 | 0.758 | 0.781 | 0.190 | 0.232 | 0.235 |
| IVW+metaIV | 0.322 | 0.296 | 0.397 | 0.160 | 0.149 | 0.135 |
| Egger | 0.256 | 0.381 | 0.669 | 0.129 | 0.110 | 0.103 |
| MVMR-IVW | 0.653 | 0.686 | 0.688 | 0.127 | 0.117 | 0.102 |
| MVMR-Egger | 0.527 | 0.565 | 0.617 | 0.142 | 0.113 | 0.132 |
| MVMR-Lasso | 0.765 | 0.799 | 0.790 | 0.202 | 0.181 | 0.136 |
| MVMR-Median | 0.686 | 0.727 | 0.855 | 0.158 | 0.166 | 0.191 |
| MVMR-Robust | 0.462 | 0.514 | 0.512 | 0.058 | 0.062 | 0.049 |
| MVcML | 0.202 | 0.309 | 0.469 | 0.024 | 0.043 | 0.058 |

Table S4: Simulation results evaluating the performance of mintMR versus its variants and competing methods when the number of samples for each exposure varies from 500 to 10,000. Sample size for outcome is fixed at 50,000. Two exposures are generated and each exposure has 5 tissues. The number of IVs is 15. The causal effects are generated from $\mathcal{N}(0, 0.02)$ and the proportion of the variance in the outcome explained by UHP effects is 0.1. Results are underlined for methods unable to control type I error rates ($\geq 0.1$).

| | Variance for generating causal effect | | | | | |
|---|---|---|---|---|---|---|
| | 0.005 | 0.01 | 0.02 | 0.005 | 0.01 | 0.02 |
| | Power | | | Type I error rate | | |
| mintMR | 0.548 | 0.734 | 0.815 | 0.050 | 0.049 | 0.050 |
| $\text{mintMR}_{\text{oracle}}$ | 0.584 | 0.764 | 0.865 | 0.050 | 0.049 | 0.050 |
| $\text{mintMR}_{\text{single-gene}}$ | 0.417 | 0.547 | 0.694 | 0.113 | 0.115 | 0.120 |
| IVW+metaIV | 0.307 | 0.351 | 0.370 | 0.136 | 0.145 | 0.158 |
| Egger | 0.214 | 0.236 | 0.271 | 0.131 | 0.134 | 0.131 |
| MVMR-IVW | 0.296 | 0.444 | 0.638 | 0.122 | 0.122 | 0.122 |
| MVMR-Egger | 0.276 | 0.383 | 0.508 | 0.124 | 0.122 | 0.126 |
| MVMR-Lasso | 0.415 | 0.631 | 0.825 | 0.177 | 0.194 | 0.211 |
| MVMR-Median | 0.382 | 0.526 | 0.700 | 0.117 | 0.114 | 0.110 |
| MVMR-Robust | 0.178 | 0.276 | 0.430 | 0.070 | 0.066 | 0.064 |

Table S5: Simulation results evaluating the performance of mintMR versus its variants and competing methods. Causal effects $\beta_{gk,l}$'s are generated from $\mathcal{N}\left(0, \sigma_\beta^2\right)$ and $\sigma_\beta^2$ varies from 0.005 to 0.02. The proportion of the variance in the outcome explained by UHP effects is 0.1. Results are underlined for methods unable to control type I error rates ($\geq 0.1$).

| Method | Tissue sample size in one tissue | | | Variance for generating causal effect | | |
|---|---|---|---|---|---|---|
| | 500 | 1000 | 10000 | 0.005 | 0.01 | 0.02 |
| mintMR | 0.032 | 0.025 | 0.017 | 0.030 | 0.032 | 0.035 |
| $\text{mintMR}_{\text{oracle}}$ | 0.015 | 0.013 | 0.012 | 0.014 | 0.015 | 0.018 |
| $\text{mintMR}_{\text{single-gene}}$ | 0.072 | 0.051 | 0.034 | 0.064 | 0.072 | 0.081 |
| IVW+metaIV | 0.048 | 0.046 | 0.028 | 0.042 | 0.048 | 0.059 |
| Egger | 0.178 | 0.142 | 0.138 | 0.162 | 0.178 | 0.206 |
| MVMR-IVW | 0.055 | 0.029 | 0.017 | 0.053 | 0.055 | 0.060 |
| MVMR-Egger | 0.088 | 0.033 | 0.019 | 0.084 | 0.088 | 0.094 |
| MVMR-Lasso | 0.063 | 0.029 | 0.017 | 0.058 | 0.063 | 0.070 |
| MVMR-Median | 0.063 | 0.034 | 0.020 | 0.060 | 0.063 | 0.068 |
| MVMR-Robust | 0.055 | 0.029 | 0.017 | 0.053 | 0.055 | 0.060 |
| MVcML | 0.036 | 0.031 | 0.022 | 0.034 | 0.036 | 0.037 |

Table S6: RMSE comparison of mintMR versus its variants and competing methods when varying the number of samples in each tissue and effect size. The proportion of the variance in the outcome explained by UHP effect is 0.1. When varying sample sizes of exposures from 500 to 10,000, the causal effects are generated from $\mathcal{N}(0, 0.02)$. When varying the variance for generating causal effects ($\sigma_\beta^2$), the sample size of exposure was fixed at 500.

| | Proportion of the variance in the outcome explained by UHP effects | | | | | |
|---|---|---|---|---|---|---|
| | 0.05 | 0.1 | 0.15 | 0.05 | 0.1 | 0.15 |
| | Power | | | Type I error rate | | |
| mintMR | 0.843 | 0.786 | 0.724 | 0.053 | 0.050 | 0.052 |
| $\text{mintMR}_{\text{oracle}}$ | 0.915 | 0.869 | 0.814 | 0.052 | 0.053 | 0.048 |
| $\text{mintMR}_{\text{single-gene}}$ | 0.644 | 0.627 | <u>0.599</u> | 0.062 | 0.098 | <u>0.126</u> |
| IVW+metaIV | <u>0.399</u> | <u>0.407</u> | <u>0.407</u> | <u>0.216</u> | <u>0.234</u> | <u>0.234</u> |
| Egger | 0.398 | 0.342 | 0.308 | 0.084 | 0.083 | 0.085 |
| MVMR-IVW | 0.673 | 0.608 | 0.538 | 0.097 | 0.094 | 0.090 |
| MVMR-Egger | <u>0.572</u> | <u>0.535</u> | <u>0.490</u> | <u>0.118</u> | <u>0.121</u> | <u>0.120</u> |
| MVMR-Lasso | <u>0.732</u> | <u>0.710</u> | <u>0.690</u> | <u>0.137</u> | <u>0.153</u> | <u>0.186</u> |
| MVMR-Median | 0.693 | 0.616 | 0.532 | 0.064 | 0.082 | 0.092 |
| MVMR-Robust | 0.508 | 0.430 | 0.373 | 0.045 | 0.046 | 0.044 |

Table S7: Simulation results evaluating the performance of mintMR versus its variants and competing methods when IVs are correlated with genetic correlation up to 0.5. Causal effects are generated from $\mathcal{N}(0, 0.02)$. Results are underlined for methods unable to control type I error rates ($\geq 0.1$).

| Category | Trait | # Genes analyzed | # Significant genes | # Significant CpGs |
|---|---|---|---|---|
| Skeletal | Height | 3434 | 139 | 59 |
| Metabolic | Body Mass Index | 3471 | 120 | 55 |
| Immunological | Platelet Count | 3418 | 111 | 34 |
| Immunological | Lymphocyte Count | 3418 | 110 | 45 |
| Immunological | Red Blood Cell Count | 3418 | 110 | 45 |
| Psychiatric | Schizophrenia | 3471 | 110 | 35 |
| Metabolic | Body Fat Percentage | 3471 | 109 | 30 |
| Immunological | White Blood Cell Count | 3418 | 106 | 33 |
| Cardiovascular | Atrial Fibrillation | 3486 | 104 | 43 |
| Immunological | Granulocyte Count | 3418 | 99 | 36 |
| Cognitive | Intelligence | 3471 | 99 | 35 |
| Immunological | Myeloid White Cell Count | 3418 | 98 | 33 |
| Immunological | Eosinophil Count | 3418 | 96 | 36 |
| Immunological | Monocyte Count | 3419 | 94 | 32 |
| Immunological | Sum Neutrophil Eosinophil Count | 3418 | 94 | 37 |
| Immunological | Neutrophil Count | 3418 | 93 | 34 |
| Neoplasms | Breast Cancer | 3471 | 93 | 36 |
| Immunological | Sum Basophil Neutrophil Count | 3418 | 92 | 34 |
| Neurological | Stroke | 3472 | 91 | 35 |
| Immunological | Sum Eosinophil Basophil Count | 3418 | 91 | 34 |
| Respiratory | Asthma | 3486 | 90 | 48 |
| Immunological | High Light Scatter Reticulocyte Count | 3418 | 88 | 40 |
| Immunological | Reticulocyte Count | 3418 | 88 | 32 |
| Metabolic | Birth Weight | 3471 | 87 | 31 |
| Psychiatric | Neuroticism Score | 3471 | 84 | 31 |
| Psychiatric | Morning or Evening Person | 3471 | 81 | 27 |
| Metabolic | Low-density Lipoprotein | 3387 | 79 | 30 |
| Metabolic | Intermediate-density Lipoprotein | 3487 | 77 | 32 |
| Psychiatric | Alzheimer's Disease | 3458 | 72 | 26 |
| Psychiatric | Chronotype | 3305 | 70 | 19 |
| Psychiatric | Sleep Duration | 3463 | 69 | 21 |
| Metabolic | High-density Lipoprotein | 3471 | 60 | 35 |
| Cardiovascular | Hypertension | 3471 | 57 | 14 |
| Neurological | Insomnia | 3467 | 53 | 28 |
| Psychiatric | Depressive Symptoms | 3393 | 37 | 20 |

Table S8: The 35 complex traits/diseases analyzed, their categories, and the number of genes being examined for each trait/disease. The numbers of genes/CpGs showing non-zero effects in at least two tissues for each outcome at the level of FDR$< 0.05$ are listed.

| Class | Trait | Exp. adj. DNAm | Exp no adj. |
|---|---|---|---|
| Immunological | High Light Scatter Reticulocyte Count | 1.47 | 1.98 |
| Immunological | Eosinophil Count | 1.47 | 2.01 |
| Immunological | Sum Eosinophil Basophil Count | 1.48 | 2.05 |
| Immunological | Monocyte Count | 1.49 | 2.08 |
| Immunological | Lymphocyte Count | 1.49 | 2.03 |
| Immunological | Neutrophil Count | 1.52 | 2.09 |
| Immunological | Reticulocyte Count | 1.52 | 1.99 |
| Immunological | Red Blood Cell Count | 1.55 | 2.09 |
| Immunological | Granulocyte Count | 1.56 | 2.08 |
| Immunological | White Blood Cell Count | 1.57 | 2.07 |
| Immunological | Sum Neutrophil Eosinophil Count | 1.58 | 2.02 |
| Immunological | Platelet Count | 1.58 | 2.09 |
| Immunological | Sum Basophil Neutrophil Count | 1.59 | 2.00 |
| Immunological | Myeloid White Cell Count | 1.59 | 2.04 |
| Psychiatric | Depressive Symptoms | 1.28 | 1.76 |
| Psychiatric | Sleep Duration | 1.30 | 1.84 |
| Psychiatric | Chronotype | 1.34 | 1.86 |
| Psychiatric | Morning or Evening Person | 1.37 | 1.93 |
| Psychiatric | Neuroticism Score | 1.40 | 1.87 |
| Psychiatric | Alzheimer's Disease | 1.41 | 1.78 |
| Psychiatric | Schizophrenia | 1.59 | 1.99 |
| Metabolic | High-density Lipoprotein | 1.39 | 1.76 |
| Metabolic | Intermediate-density Lipoprotein | 1.42 | 1.90 |
| Metabolic | Birth Weight | 1.45 | 2.03 |
| Metabolic | Low-density Lipoprotein | 1.47 | 1.94 |
| Metabolic | Body Fat Percentage | 1.55 | 2.15 |
| Metabolic | Body Mass Index | 1.63 | 2.29 |
| Cardiovascular | Hypertension | 1.24 | 1.89 |
| Neurological | Insomnia | 1.32 | 1.75 |
| Neurological | Stroke | 1.43 | 1.82 |
| Cardiovascular | Atrial Fibrillation | 1.51 | 1.87 |
| Neoplasms | Breast Cancer | 1.40 | 1.88 |
| Respiratory | Asthma | 1.53 | 1.97 |
| Cognitive | Intelligence | 1.54 | 1.95 |
| Skeletal | Height | 1.71 | 2.33 |

Table S9: The averaged genome-wide inflation factors across tissues for the p-values of gene expression on different outcomes with and without accounting for DNA methylation, using mintMR.

| Trait | mintMR | MVMR-Median | MVMR-Lasso | MVMR-IVW | MVMR-Robust | MVMR-Egger |
|---|---|---|---|---|---|---|
| Hypertension | 1.24 | 1.79 | 1.89 | 1.59 | 1.84 | 1.48 |
| Morning or Evening Person | 1.37 | 1.80 | 1.78 | 1.58 | 1.48 | 1.46 |
| Neuroticism Score | 1.40 | 1.59 | 1.71 | 1.58 | 1.58 | 1.48 |
| Birth Weight | 1.45 | 1.84 | 1.83 | 1.62 | 1.58 | 1.51 |
| Body Fat Percentage | 1.55 | 2.13 | 2.41 | 1.83 | 2.01 | 1.71 |
| Body Mass Index | 1.63 | 2.19 | 2.49 | 1.85 | 2.03 | 1.74 |
| Height | 1.71 | 2.26 | 2.54 | 1.93 | 2.07 | 1.75 |
| Depressive Symptoms | 1.28 | 1.57 | 1.33 | 1.39 | 1.21 | 1.30 |
| Breast Cancer | 1.40 | 1.54 | 1.43 | 1.47 | 1.42 | 1.36 |
| Eosinophil Count | 1.47 | 1.81 | 1.71 | 1.54 | 1.51 | 1.40 |
| High Light Scatter Reticulocyte Count | 1.47 | 1.76 | 1.68 | 1.54 | 1.49 | 1.39 |
| Monocyte Count | 1.49 | 1.77 | 1.71 | 1.53 | 1.49 | 1.41 |
| Neutrophil Count | 1.52 | 1.85 | 1.77 | 1.59 | 1.53 | 1.44 |
| Platelet Count | 1.58 | 1.83 | 1.85 | 1.61 | 1.62 | 1.50 |
| Schizophrenia | 1.59 | 1.80 | 1.92 | 1.67 | 1.70 | 1.54 |
| Sleep Duration | 1.30 | 1.49 | 1.33 | 1.34 | 1.22 | 1.25 |
| Chronotype | 1.34 | 1.57 | 1.45 | 1.41 | 1.20 | 1.27 |
| Low-density Lipoprotein | 1.47 | 1.72 | 1.59 | 1.50 | 1.33 | 1.39 |
| Sum Eosinophil Basophil Count | 1.48 | 1.82 | 1.68 | 1.50 | 1.48 | 1.38 |
| Lymphocyte Count | 1.49 | 1.76 | 1.74 | 1.60 | 1.47 | 1.47 |
| Reticulocyte Count | 1.52 | 1.76 | 1.73 | 1.58 | 1.49 | 1.43 |
| Asthma | 1.53 | 1.70 | 1.56 | 1.54 | 1.38 | 1.45 |
| Intelligence | 1.54 | 1.71 | 1.67 | 1.56 | 1.41 | 1.42 |
| Red Blood Cell Count | 1.55 | 1.84 | 1.82 | 1.61 | 1.52 | 1.49 |
| Granulocyte Count | 1.56 | 1.88 | 1.80 | 1.61 | 1.53 | 1.44 |
| White Blood Cell Count | 1.57 | 1.85 | 1.81 | 1.64 | 1.53 | 1.51 |
| Sum Neutrophil Eosinophil Count | 1.58 | 1.89 | 1.79 | 1.62 | 1.52 | 1.46 |
| Myeloid White Cell Count | 1.59 | 1.89 | 1.79 | 1.60 | 1.55 | 1.45 |
| Sum Basophil Neutrophil Count | 1.59 | 1.88 | 1.76 | 1.60 | 1.51 | 1.43 |
| Intermediate-density Lipoprotein | 1.42 | 1.65 | 1.45 | 1.40 | 1.27 | 1.30 |
| Insomnia | 1.32 | 1.44 | 1.27 | 1.29 | 1.11 | 1.22 |
| High-density Lipoprotein | 1.39 | 1.39 | 1.21 | 1.26 | 1.07 | 1.16 |
| Stroke | 1.43 | 1.59 | 1.36 | 1.36 | 1.22 | 1.28 |
| Atrial Fibrillation | 1.51 | 1.60 | 1.44 | 1.42 | 1.22 | 1.34 |
| Alzheimer's Disease | 1.41 | 1.38 | 1.31 | 1.32 | 1.28 | 1.20 |

Table S10: The averaged genome-wide inflation factors across tissues for the p-values of gene expression adjusting for cis-DNAm on different outcomes based on mintMR and five competing MVMR methods. Results from competing methods are underlined if the method has a higher inflation factor than mintMR.
